# Two modes of inhibitory neuronal shutdown distinctly amplify seizures in humans

**DOI:** 10.1101/2020.10.09.20204206

**Authors:** Omar J. Ahmed, Tibin T. John, Shyam K. Sudhakar, Ellen K.W. Brennan, Alcides Lorenzo Gonzalez, Jason S. Naftulin, Emad Eskandar, Joseph R. Madsen, G. Rees Cosgrove, Andrew S. Blum, N. Stevenson Potter, George A. Mashour, Leigh R. Hochberg, Sydney S. Cash

## Abstract

Inhibitory neurons are critical for normal brain function but dysregulated in disorders such as epilepsy. At least two theories exist for how inhibition may acutely decrease during a seizure: hyperpolarization of fast-spiking (FS) inhibitory neurons by other inhibitory neurons, or depolarization block (DB) of FS neurons resulting in an inability to fire action potentials. Firing rate alone is unable to disambiguate these alternatives. Here, we show that human FS neurons can stop firing due to both hyperpolarization and DB within the same seizure. However, only DB of FS cells is associated with dramatic increases in local seizure amplitude, unobstructed traveling waves, and transient increases in excitatory neuronal firing. This result is independent of seizure etiology or focus. Computational models of DB reproduce the *in vivo* human biophysics. These methods enable intracellular decoding using only extracellular recordings in humans and explain the otherwise ambiguous inhibitory neuronal control of human seizures.

## INTRODUCTION

Epilepsy is a debilitating disease affecting some 50 million people worldwide^1,2^. Epileptic seizures are thought to result from an imbalance between excitatory and inhibitory neuronal activity^3^. However, electrographically similar seizures on the macroscopic scale can be driven by mechanistically distinct processes on the cellular and cell-type population scale^4^. Thus, the relative roles of local inhibitory and excitatory networks in driving seizure progression must be elucidated to guide the development of novel treatments for intractable epilepsies and our understanding of their associated seizures. Due to the tremendous technical challenges of recording from individual human neurons and the relative sparsity of inhibitory neurons, the activity of well isolated inhibitory interneurons during seizures is only rarely examined^5,6^.

There are several theories, based on slice and whole animal experiments, proposing both insufficient^7–15^ and excessive^16–20^ inhibition as possible facilitators of epileptic activity, with at least two hypotheses for how inhibition may acutely decrease during a seizure: (1) hyperpolarization of fast-spiking (FS) inhibitory neurons by other inhibitory neurons^21–24^, or (2) excessive depolarization of FS neurons that precludes subsequent action potentials due to blockade of voltage-dependent sodium channels^12,14,25,56^. Resolving these two theories necessitates whole cell recordings^26^, but it is practically impossible to record the intracellular membrane potential of neurons during human seizures *in vivo*. In this study, we address these challenges by combining (1) large-scale extracellular recordings of human neocortical inhibitory and excitatory neurons during focal seizures with secondary generalization and (2) a novel method of decoding membrane potential trajectory from extracellular action potentials. We show the remarkable ability of active fast-spiking inhibitory neurons to block epileptic traveling waves in human neocortex, and reveal the dynamical control of human seizures by the subthreshold trajectory of inhibitory neurons.

## RESULTS

Patients were implanted with intracranial grid electrodes as part of the clinical process of identifying the precise site of origin of their drug-resistant focal epilepsy (see Materials and Methods). A 4×4 mm NeuroPort microarray (Blackrock Microsystems) was also placed in a region of the neocortex that was expected to be in the resection site (Figure 1a; Extended Data Fig. 1a). Histology of the resected tissue confirmed that the electrodes consistently targeted layers 2/3 of the neocortex (Extended Data Fig. 1b). We used these arrays to simultaneously record the activity of dozens of individual neocortical neurons during both ictal and interictal activity. We then classified the neurons as either fast-spiking (FS) inhibitory interneurons or regular-spiking (RS) excitatory cells using well-established criteria^27,28^, including action potential shape (Extended Data Fig. 1c-f). FS cells correspond to the class of parvalbumin-expressing interneurons and represent the largest source of inhibition in the neocortex^29^. The resulting information thus allowed us to differentiate between putative inhibitory and excitatory unit activity patterns during seizure progression in humans.

**Figure 1:**
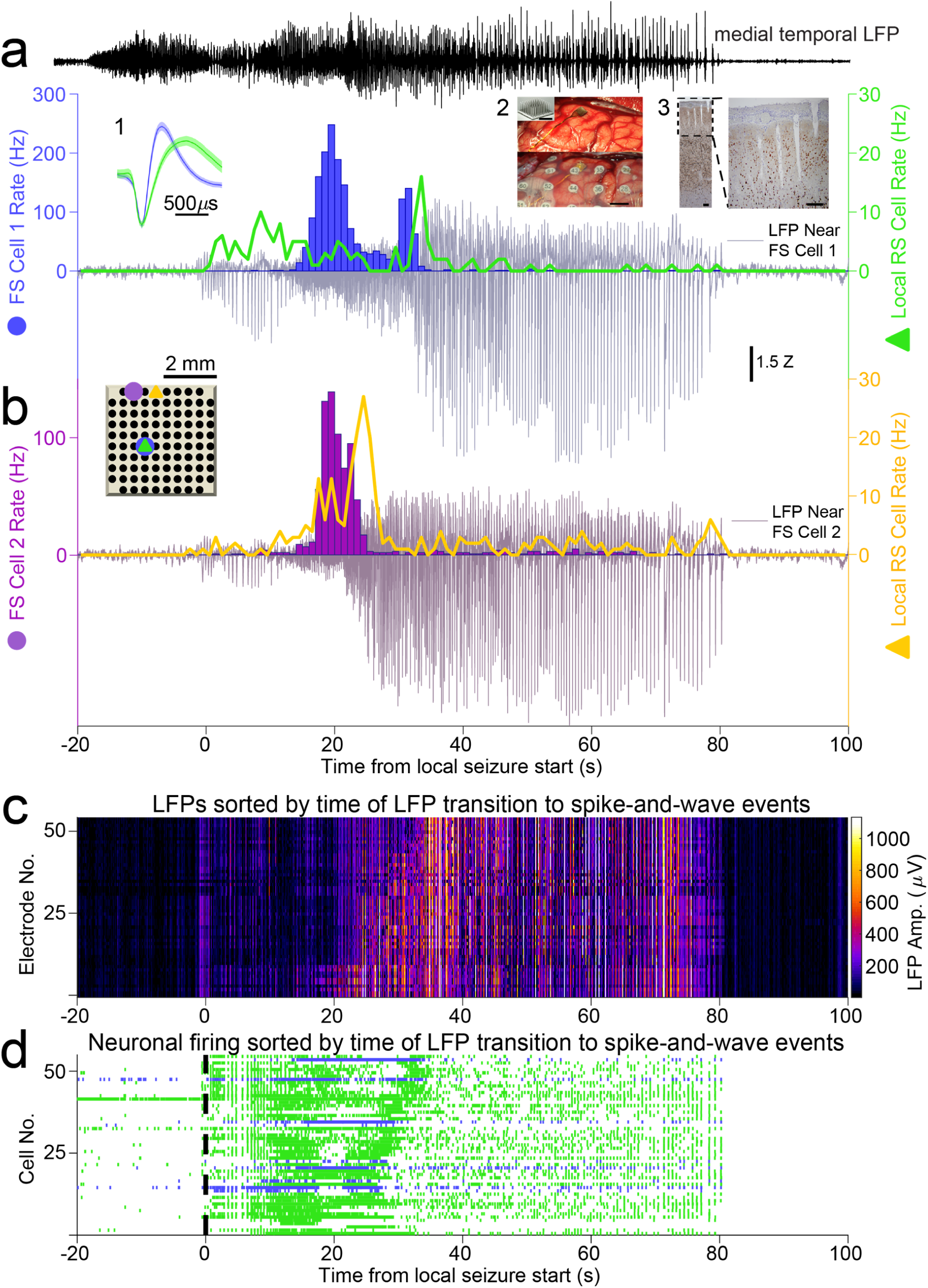
Human neocortical inhibitory and excitatory neurons have different temporal profiles relative to secondarily generalized focal seizure progression on local electrodes. **a. *Upper***. Electrocorticogram trace (ECoG, black) from contact closest to source of focal seizure in hippocampus of Patient A. Note its time of initiation at T = -20 seconds, substantially preceding the start of the seizure in neocortex.***Lower***. The firing rate of a single FS cell (blue), the firing rate of a single RS cell recorded from the same electrode (green) and the local field potential (LFP, gray, recorded from the same electrode as the FS cell) during a seizure recorded from Patient A. Note the decrease in LFP amplitude as the FS cell starts firing, followed by a dramatic increase in LFP seizure amplitude after the FS cell decreases its rate at T=35 seconds. RS cell quickly increases its firing rate as the seizure first spreads to the NeuroPort array (T=0 seconds), then decreases in rate slightly as the FS cell switches on, and finally settles into a lower firing rate regime during the large spike-and-wave events that dominates the remainder of the seizure after FS cell cessation. Inset shows the mean normalized extracellular action potential waveforms (with 99% confidence intervals in lighter shading) of FS Cell 1 (blue) and the neighboring RS cell (green). Inset 1: the mean normalized extracellular action potential waveforms (with 99% confidence intervals in lighter shading) of FS Cell 1 (blue) and the neighboring RS cell (green). Inset 2: The implanted microelectrode array (Top) with overlaid clinical grid electrodes (Bottom); Scale bar: 1 cm. Top left corner shows the array before implantation. Scale bar: 200 µm. Inset 3: Histology of resected tissue showing the NeuN-stained neurons in the full neocortical column and electrode tracts and an enlarged image (Right) showing that the array targeted layer 3 of the neocortex. Scale bars: 400 µm. **b**. The inset shows a schematic of the array and the location of two simultaneously recorded FS cells separated by ∼2 mm: FS Cell 1 (blue, firing rate shown in Fig. **1c**) and FS Cell 2 (purple). FS Cell 2 approached cessation at T=24 seconds, well before FS Cell 1. The LFP (grayish-purple) recorded at the same location as FS Cell 2 dramatically increases at the same time as FS Cell 2 decreases firing, well before the increase seen in the LFP recorded next to FS Cell 1. These suggest the activity of FS cells during human seizures is strongly correlated to the local intensity of seizure waves. FS Cell 2 activity cessation again precedes a sharp increase in local RS cell activity (gold), further suggesting an important role of FS cells in controlling local activity during seizure progression. **c**. Heatmap shows local LFP amplitude (absolute value) over time on each electrode in NeuroPort array exhibiting classifiable units as each row, sorted by time of start of spike-and-wave event and with brighter colors indicating larger amplitudes. Note the increasing delay to start of spike- and-wave event suggesting different dynamics to seizure spread across the array over the timecourse of approximately 20 seconds. **d**. Raster plot showing spike times of all cells on NeuroPort array in Patient A that could be classified into FS (blue) or RS (green) categories with rows sorted by the same order as in (C). Note the increasing delay to reduction in spike density corresponding to LFP transition to spike- and-wave events suggesting control of local seizure progression by local cellular spiking activity.

A total of 37 FS cells and 539 RS cells were recorded across 4 patients with NeuroPort arrays implanted in the temporal neocortex (see Supplementary Methods for details about each patient). As secondarily generalized seizures first reached the arrays, both FS and RS cells increased their rate of action potentials (Figs. 1, 2, and Extended Data Figs. 2-4). This finding is consistent with what is known about the feedforward recruitment of both inhibition and excitation^25,30^. At the population level, the FS firing rate was significantly higher than RS cells (100,000 bootstrap iterations of label-shuffled peak rate differences; p=0.036). Within 40 seconds of seizure onset, the mean FS firing rate fell rapidly to 0-2 Hz at the same time as local field potential intensity increased dramatically to its highest levels (Figs. 1, 2, and Extended Data Figs. 2-4). Consistently, among the best isolated FS units during the seizure (isolation quality discussed in more detail below and see Methods), 10 out of 15 had at least one nearby RS cell that exhibited the peak of its overall activity several seconds after the dramatic fall in local FS activity and accompanying elevation in local field potential intensity (Figs. 1, 2, and Extended Data Figs. 2-4). Thus, at the population level, FS cells show a cessation of activity near the middle of secondarily generalized seizures accompanied by a dramatic increase in the amplitude of seizure activity. This FS cessation is followed by a transient increase in RS cell firing, presumably because these RS cells are now less inhibited due to the loss of FS firing. The RS population rate eventually also fell to an average of 0-3 Hz but with a delay of several seconds following FS cells. At the population level, FS cells (N=37) exhibited significantly earlier cessation times than RS cells (N=539) (Fig. 2g-j, FS cell average cessation occurred 4.6 seconds earlier than RS average cessation; 100,000 bootstrap iterations of label-shuffled time differences; p<0.0001).

**Figure 2:**
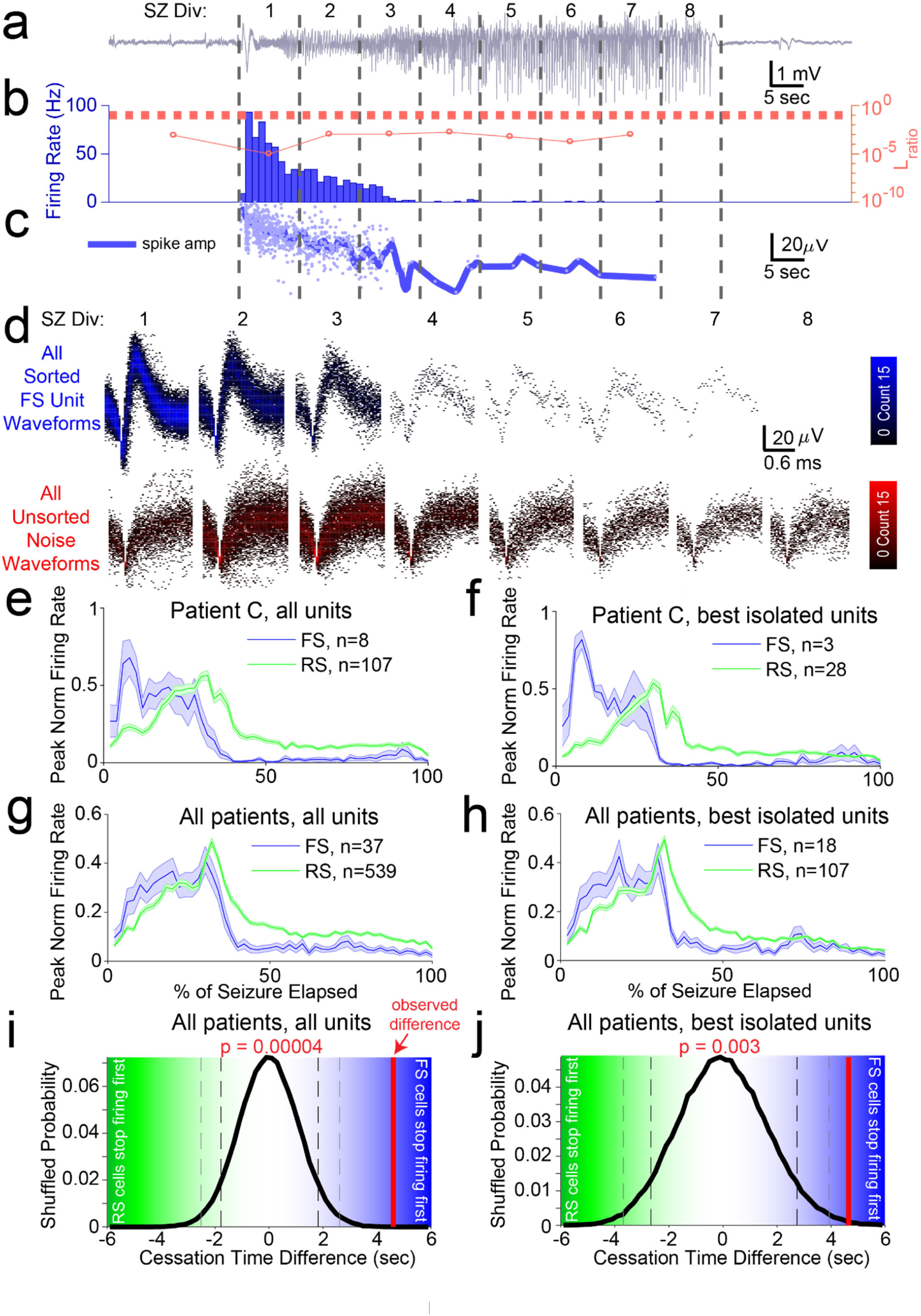
Analyzing unit subpopulations by isolation quality demonstrates consistent cell-type specific activity profiles and cessation order despite changing unit amplitude and noise structure. **a**. LFP in Patient C indicating seizure as split into 8 equal time divisions for analysis of unit isolation quality across duration of seizure. **b**. Bar graph shows firing rate in 1 second bins of best-isolated example FS unit (blue) in Patient C. Dotted red line indicates threshold used to determine best-isolated units using the dynamic L_ratio_ measure (see Methods) in each time division of seizure. Line plot indicates dynamic L_ratio_ in each division (red) and shows large separation of example FS unit from noise in feature space used for clustering throughout the seizure (note log scale). **c**. Line plot showing average spike amplitude (dark blue) and individual spike amplitudes (light blue) of example FS unit over course of seizure in Patient C. Note that even as amplitude decreases the unit remains well-isolated from noise as quantified by dynamic L_ratio_ across seizure. **d**. Time-voltage histogram of all threshold crossings assigned to this example FS unit (blue, *Upper)* and to noise (red, *Lower*) in eight divisions of seizure in Patient C. Shows unit waveforms are visually distinguishable from threshold crossings assigned as noise across seizure. **e**. Average firing rate traces across seizures for each cell type group in Patient C, with y-axis normalized to firing rate peak and x-axis normalized to total seizure duration before averaging. Note right-shifted rate profile of RS units over course of seizure as compared to FS units. **f**. Average firing rate traces across seizures for each cell type group using only best-isolated units (see Methods) in Patient C, with y-axis normalized to firing rate peak and x-axis normalized to total seizure duration before averaging. Note right-shifted rate profile of best-isolated RS units over course of seizure as compared to best-isolated FS units. **g**. Average firing rate traces across seizures for each cell type group across all patients, with y-axis normalized to firing rate peak and x-axis normalized to total seizure duration before averaging. Note right-shifted rate profile of RS units over course of seizure as compared to FS units. **h**. Average firing rate traces across seizures for each cell type group using only best-isolated units (see Methods) across all patients, with y-axis normalized to firing rate peak and x-axis normalized to total seizure duration before averaging. Note right-shifted rate profile of best-isolated RS-classified units over course of seizure as compared to best-isolated FS-classified units. **i**. Observed time difference (solid red line) between mean RS activity cessation timing (n=399) and mean FS activity cessation timing (n=37) in relation to probability distribution of this timing difference across random label reshufflings of unit cell type labels (N_shuffles_=100,000; solid black line). P-value indicates probability of observing a value equal to or more extreme than the observed value simply by random assignment of cell type to each unit. Values on the right half correspond to a positive difference between RS and FS cessation, with larger values associated with increasing certainty that FS cells stop firing before RS cells. Values on the left half correspond to a negative difference between RS and FS cessation times, with larger values associated with increasing certainty that RS cells stop firing before FS cells. **j**. Observed time difference (solid red line) between mean best-isolated RS activity cessation timing (n=83; Lratio < 0.1 during middle 3 seizure divisions) and mean best-isolated FS activity cessation timing (n=17; Lratio < 0.1 during middle 3 seizure divisions) in relation to probability distribution of this timing difference across random label reshufflings of unit cell type labels (N_shuffles_=100,000; solid black line). P-value indicates probability of observing a value equal to or more extreme than the observed value simply by random assignment of cell type to each unit. Values on the right half correspond to a positive difference between best-isolated RS and best-isolated FS cessation times, with larger values associated with increasing certainty that best-isolated FS cells stop firing before best-isolated RS cells. Values on the left half correspond to a negative difference between best-isolated RS and best-isolated FS cessation, with larger values associated with increasing certainty that best-isolated RS cells stop firing before best-isolated FS cells.

We next compared activity patterns among individual FS cells. Many individual FS cells did not start firing until ∼10 seconds into the seizure, often starting to fire robustly only after the RS rate had already increased (Figs. 1, 2, and Extended Data Figs. 2-4). As seen with the population means (Fig. 2g), each individual FS cell dramatically reduced its firing roughly half-way through the seizure (Figs. 1, 2, and Extended Data Figs. 2-4). However, even simultaneously recorded FS cells during a single seizure did not necessarily cease firing at the same time, just as they did not start firing at the same time. Figure 1 shows the activity of two simultaneously recorded FS cells that were separated by a distance of 2.04 mm. FS Cell 1 (Fig. 1a) approached cessation ∼35 seconds into the seizure, and at ∼35 seconds, the amplitude of the LFP next to FS Cell 1 increased dramatically, as did the firing rate of nearby RS cells. FS Cell 2 (Fig. 1b) approached cessation ∼24 seconds into the seizure, and the LFP amplitude next to FS Cell 2 also increased at ∼24 seconds, along with the firing rate of nearby RS cells. Furthermore, when sorting all simultaneously recorded channels during a seizure by the timing of transition to the large amplitude spike-and-wave event phase, a raster plot of all spikes with units sorted in the same order produced a similar sequence of mid-seizure firing time cessations (Fig. 1c&d). Thus, FS cell cessation is correlated with a dramatic increase in the very local intensity of seizures.

Given changing waveform shapes and noise characteristics during seizures^31^, an important methodological question arises of how separable clusters in waveform feature space are from each other and from noise to allow valid unit assignments during this period. To quantitatively assess this isolation quality of feature space clusters over the course of seizures, we employed a modified version of the L_ratio_ metric of cluster separation introduced and validated by Schmitzer-Torbert et al. (2005)^58^ (see Methods). Cluster divisions with L_ratio_ less than 0.1 are considered to have a significantly low level of false negative contamination, indicating that drops in firing rate are not caused by over-assigning candidate spikes to noise or other clusters due to changing waveform shapes in these divisions. We consider units whose average contamination level across seizure divisions (dynamic L_ratio_) is less than 0.1 to be a best-isolated subset^58^. Figure 2 shows an example of an FS unit whose waveform shape could be well isolated from noise across the entire seizure despite showing a monotonic decrease in spike amplitude (Fig. 2a-c). The isolation quality of this unit throughout the seizure is demonstrated visually by 2D voltage histograms of these waveforms (Fig. 2d) and is quantitatively captured by its dynamic L_ratio_ being far below threshold in all seizure divisions in which it was active (Fig. 2b). This isolation quality was similarly sustainable throughout the seizure for many RS units (e.g. Extended Data Fig. 5), although at a lower proportion than FS units, with 48.6% of FS units meeting this criterion but only 19.8% of RS units. This pattern sustained throughout seizures, with the proportion of well-isolated FS units remaining between 30% and 48% across all 8 seizure divisions and that of RS units remaining between 13% and 19% (Extended Data Fig. 6). The apparent difference in sortability by cell type may be because the frequency content of noise is such that random threshold crossings are more similar in shape and amplitude to the wider waveforms of RS units than to the sharper waveforms of FS units. This suggests that FS cell spikes are inherently more distinguishable from noise in human neocortical recordings even during seizures. We considered the best-isolated subset of units following this quantitative criterion in parallel with all-inclusive analyses of firing rate dynamics in various time divisions of seizures with similar results. In particular, similar temporal profiles by cell type were observed throughout seizures when considering all units or only best-sorted units with an average L_ratio_ < 0.1 across all divisions for Patient C (Fig. 2e&f), as well as when comparing average firing rate time courses by cell type across all patients (Fig. 2g&h). Furthermore, the observed time difference between mean RS activity cessation timing and mean FS activity cessation timing remained significantly larger than that expected by random cell-type label reshuffling when considering all units (p<0.0001; Fig. 2i), as well as only best-isolated units with maximum L_ratio_ < 0.1 (p<0.005; Fig. 2j). Thus, FS cell cessation occurs several seconds before RS cell cessation across the population as well as for the very best isolated units.

The firing profile of individual FS cells and the accompanying changes in the seizure LFP amplitude were remarkably consistent across all secondary generalized focal seizures examined, independent of etiology or focus (Figs. 1, 2, and Extended Data Figs. 2-4). In patients with an etiology of mesial temporal sclerosis, where the seizure started focally in the hippocampus or surrounding medial temporal regions, FS cells in the temporal neocortex consistently stopped firing after the seizure secondarily generalized and spread to the neocortex, leading to accompanying increases in seizure LFP amplitude (Fig. 1a&b). Similarly, in a patient with an etiology of neocortical dysplasia (Patient C) where the seizure originated in the temporal neocortex, several cm from the implanted electrode array, FS cells near the array stopped firing within the first 40 seconds of the seizure spreading to that location (Fig. 2a&b). Again, this coincided with a transition to even larger amplitude spike-wave LFP events.

The variability in FS cessation times raises the question of how seizure waves travel across the neocortex: is there a wave that slowly moves across the cortex^32^ or a series of faster waves that are perhaps altered by the local activity of FS cells^11,30,33^? We found fast traveling LFP waves in all patients that swept across the 4×4 mm microelectrode array within 40 ms, at a speed of ∼0.1 m/s, consistent with estimates from slice and computational studies^30,34–36^. There was a dramatic effect of individual FS cell activity: when an FS cell was still firing, it was able to impede and alter epileptic traveling waves (Fig. 3a), preventing the wave from increasing the seizure’s LFP amplitude in the vicinity of the FS cell. Once the FS cell activity ceased, traveling waves swept through the entire array, successfully recruiting the area around the now-silent FS cell (Figure 3B). Indeed, once all FS cells had switched off, the path of epileptic traveling waves became far more regular and stereotyped. Thus, human FS inhibitory cells possess a remarkable capability to obstruct and alter the path of epileptic traveling waves. This again points to the importance of FS activity in controlling local neuronal activity and LFP dynamics during local seizure propagation but leaves open the question of what is causing them to stop firing during the seizure.

**Figure 3:**
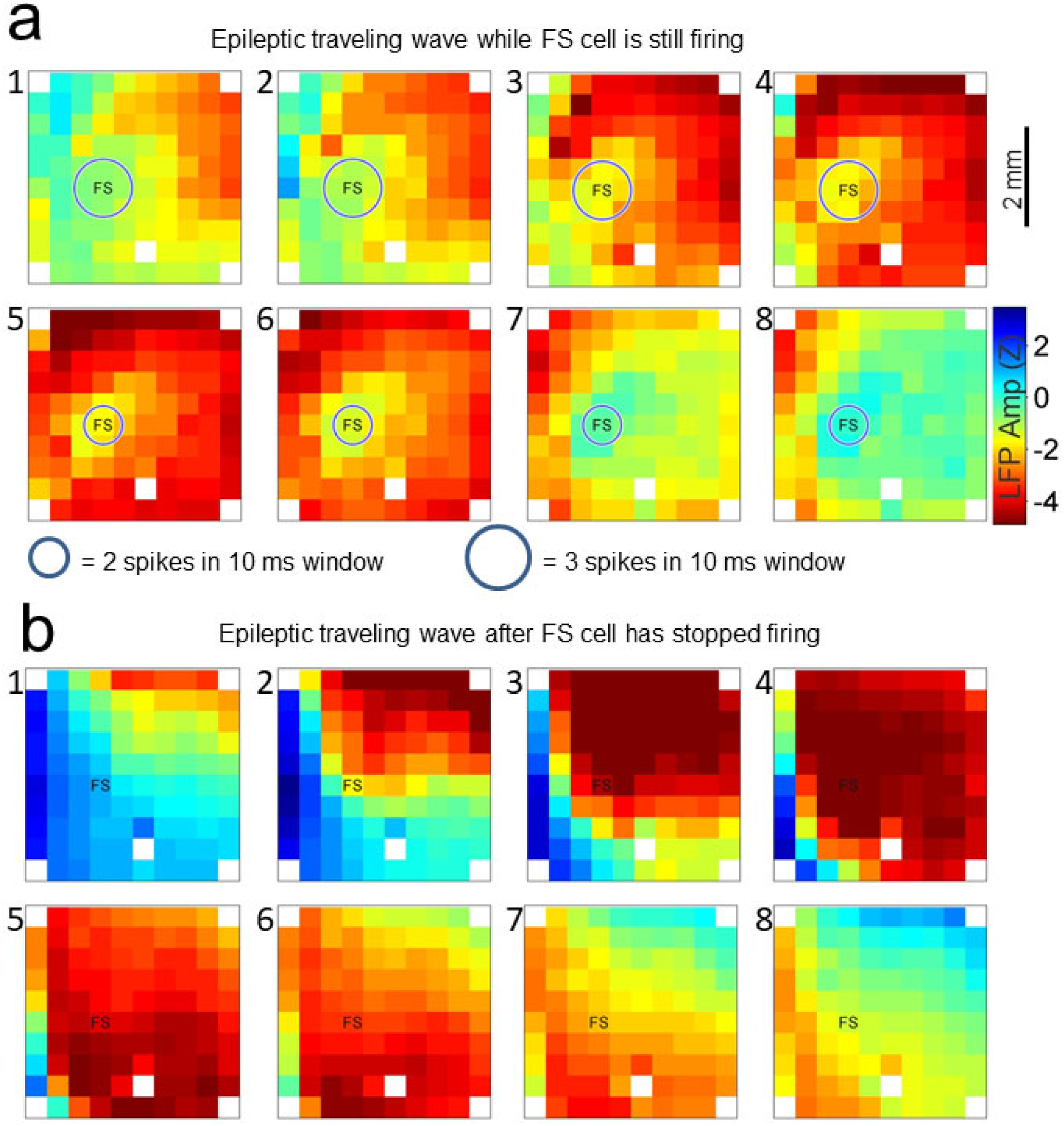
Fast-spiking inhibitory neurons, when still firing, impede the spread of epileptic traveling waves. **a**. An example of a traveling wave recorded 30 seconds after the start of a seizure in Patient A. The FS cell located at the position marked “FS” was firing at high rates at this time. The 8 snapshots (1-8) are taken over a 35 ms interval as the traveling LFP wave starts in the top-right corner of the array and travels across the array. The traveling wave does not fully invade the region of neocortex containing the active FS cell, resulting in lower amplitude LFPs around the FS cell. In each subfigure, each square denotes a single LFP sensor, and white squares indicate omitted sensors. **b**. An example of a traveling wave recorded 49 seconds after the start of the same seizure. In the absence of FS cell firing, the epileptic traveling wave moves unimpeded through the array, fully invading the region around the marked FS cell and resulting in larger amplitude LFPs at all locations.

There are two ways in which FS cells can stop firing during a seizure: 1) they could be hyperpolarized by inhibition from other inhibitory neurons^22^, also known as the disinhibition hypothesis^23^; or 2) they could enter depolarization block and thus become incapable of firing additional action potentials, despite receiving strong excitatory synaptic input^12,14,25,56^. These two scenarios produce very different predictions for how the action potential (AP) amplitude of an FS cell should change during a seizure. In the case of hyperpolarization, the AP amplitude should increase as the firing rate is decreasing. In the case of depolarization block, AP amplitude should decrease before the cell stops firing while its firing rate is also decreasing. To monitor the relative contributions of hyperpolarization and excessive depolarization throughout the seizure in each recorded unit, we therefore devised a novel method that decodes the membrane potential regime of neurons from extracellular spike amplitude data by computing the sign of the correlation between AP amplitude and firing rate in each second of the seizure (Fig. 4a-c; see Methods). Some units first paused their firing due to transient inhibition from other cells, as suggested by strongly negative correlations between AP amplitude and firing rate as firing rate initially decreased (Fig. 4b&h; Extended Data Figs. 7&8). This period corresponded precisely to the time period of increased firing in neighboring FS units (Fig. 4d&j). However, these apparently hyperpolarized units resumed their firing after this pause. Cessation of firing subsequently occurred presumably due to massive depolarization of their membrane potential, as demonstrated by strongly positive correlations of AP amplitude and firing rate as these units stopped firing spikes (Fig. 4b&h). The regimes of negative correlation correspond to a dynamic inhibitory control of local firing rates during the seizure, followed by a regime of positive correlation with decreasing AP amplitudes during the final descent in firing rate, a pattern consistent with what is seen during depolarization block.

**Figure 4.**
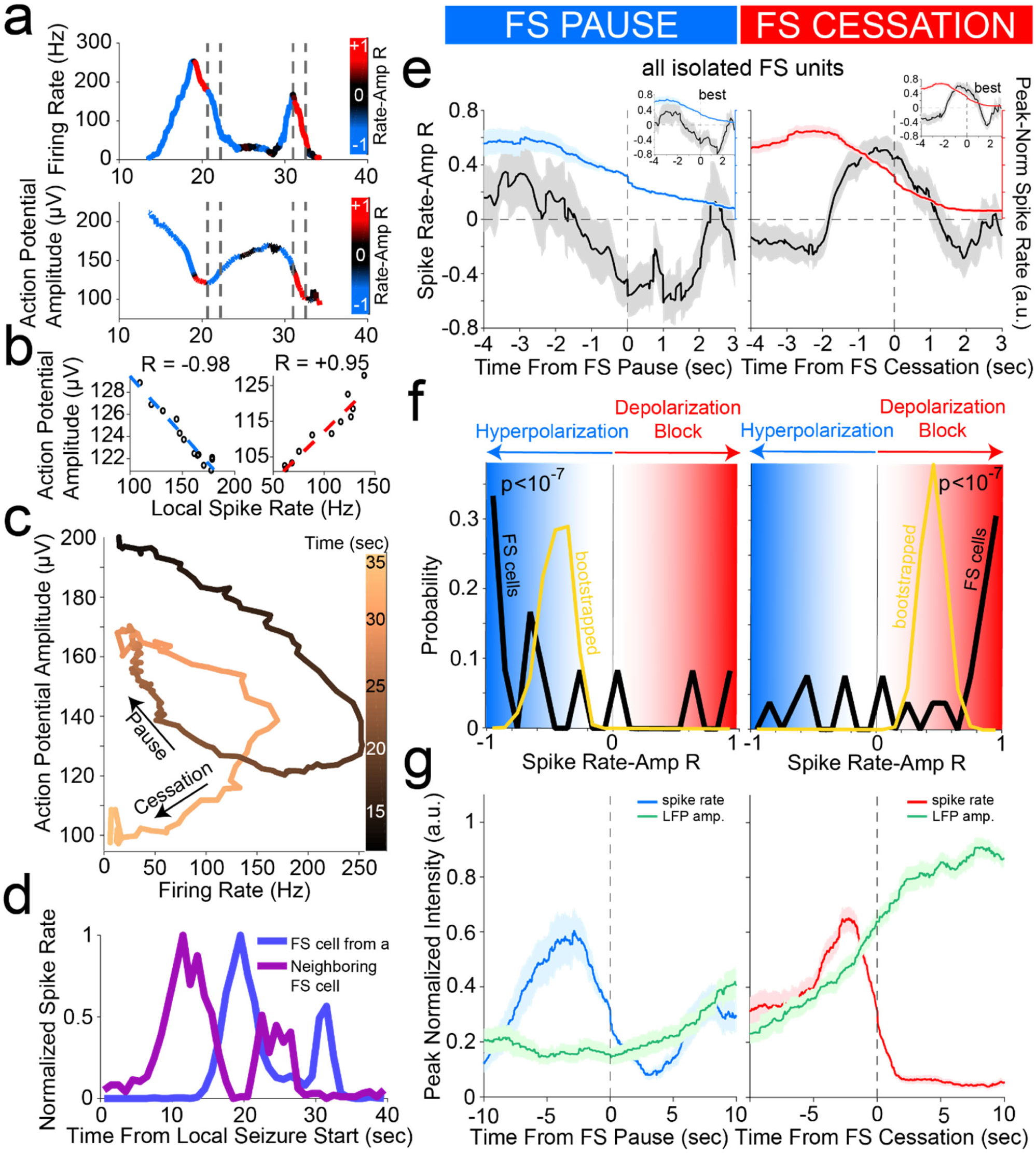
Cessation of individual FS unit activity is consistently associated with inferred membrane potential signatures of depolarization block, despite prior hyperpolarized pauses. **a**. Firing rate *(upper)* and trough-to-peak spike amplitude (*lower*) of example FS unit in Patient A, color-coded by the local correlation between spike rate and amplitude (in 1 second time bins) as an extracellular proxy for membrane potential trajectory and subthreshold input history. Dotted lines indicate starts and ends of two time periods of firing rate suppression characterized by different membrane potential signatures further characterized in (B), namely a negative correlation regime corresponding to inhibition followed by a positive correlation regime corresponding to over-excitation ending in firing rate cessation putatively though depolarization block. **b**. Example of negative correlation (*left*) between local spike rate and amplitude in first time period indicated by dotted lines in (A) and example of positive correlation (*right*) between local spike rate and amplitude in second time period indicated by dotted lines in (A). Least squares linear fit indicated in dotted lines following color scheme in (A) with Pearson’s correlation coefficient indicated above each plot. **c**. Trajectory of unit activity over time during seizure in local spike rate vs spike amplitude space, with increasing time indicated by increasingly lighter copper color. The first time period of firing rate reduction in dotted lines in (A) is indicated with an arrow as “Pause” and the second time period of firing rate reduction in dotted lines in (A) is indicated with an arrow as “Cessation.” **d**. Firing rate of unit from (A) with neighboring FS unit firing rate overlaid, giving further evidence that first period of firing rate suppression corresponds to inhibition from local FS units while second period of firing rate suppression does not correspond to inhibition from local FS units. **e**. Unit cessation-triggered population average of the time course of novel membrane potential regime measure, i.e. the correlation coefficient relating spiking amplitude and rate in a local time window, around the two significant descents in firing rate (below 30% of peak rate) that occur in sequence during seizure. These are designated as pause (*left panel*) and cessation (*right panel*). Left panel shows population average firing rate (blue) and inferred membrane potential regime (black) aligned according to the time of pause in each unit, for all FS units displaying a pause (n=14), with inset showing average for best-isolated FS units displaying a pause (n=10). Right panel shows population average firing rate (red) and inferred membrane potential regime (black) aligned according to the cessation time of each unit, for all FS units (n=37), with inset showing average for best-isolated FS units (n=24). **f**. Probability distribution of membrane potential regime measure (spike rate-amplitude correlation; black line) at the time of pause (*left panel*) for FS unit pausing subpopulation (n=14) with sample mean distribution (gold line; resampling size *n*=10, *N*_*bootstrap*_=50,000) showing the mean correlation to be significantly below zero, i.e. in the hyperpolarized membrane potential regime (blue). In combination with its declining firing rate, this is indicative of widespread inhibition across this subpopulation at the time of pause. The population distribution of inferred membrane potential regime is also shown at the time of cessation (*right panel*) for the full FS unit population (n=37) with sample mean distribution (gold line; resampling size *n*=10, *N*_*bootstrap*_=50,000) showing the mean correlation to be significantly above zero, i.e. in the highly depolarized membrane potential regime (red). This is indicative of widespread depolarization block occurring across FS population at the time of cessation. **g**. Unit cessation-triggered population average of the time course of same-electrode LFP amplitude around unit activity pause (*left panel*) and cessation (*right panel*). In particular, left panel shows population average firing rate (blue) and LFP amplitude (black) aligned according to the time of pause in each unit, for all FS units displaying a pause (n=14). Right panel shows population average firing rate (red) and LFP amplitude (black) aligned according to the time of cessation in each unit, for all FS units (n=37), indicative of larger LFP increase associated with second FS firing rate descent (cessation) than with first (pause) despite comparable local firing rates in the two conditions.

This pattern was also seen on the population level in spiking event-triggered averages of the time course of our predicted membrane potential measure (Fig. 4e). Aligning inferred membrane potential time courses around the time of firing rate pauses (dropping below 30% of peak rate) revealed a large negative deflection for FS cells, indicative of these units being inhibited via hyperpolarization to reduce firing rate at this time (Fig. 4e; n=12; R_mean,pause_=-0.44±0.19 [sem]; less than 0 with p<10^−7^ by bootstrap mean resampling test, n_sample_=10, N_bootstrap_=50,000). Furthermore, the inferred membrane potential amongst the FS cell population showed that this shift was also widespread (Fig. 4f), with 9 out of 12 (75%) FS units pausing during the negative, hyperpolarized regime during this rate-based event. Aligning inferred membrane potential time courses around the later time of firing rate cessation in units (defined by rate falling below 30% of peak rate for the final time) revealed a large positive deflection for FS cells, indicative of these units entering a regime depolarization block (Fig. 4e; n=26; R_mean,cess_=+0.45±0.12 [sem]; greater than 0 with p<10^−7^ by bootstrap mean resampling test, n_sample_=20, N_bootstrap_=50,000). Again, the inferred membrane potential amongst the FS cell population showed that this shift was also widespread across units (Fig. 4f), with 20 out of 26 (77%) FS units landing in the positive, over-excited regime. When comparing these values only in units that showed both a pause and cessation to control of heterogeneity amongst different units, the inferred membrane potential was still significantly higher as the unit ceased firing compared to as the unit paused its firing (right-sided Wilcoxon signed rank test; n=12, p=0.0049). Finally, these firing rate descents in FS cell activity were associated with distinct changes in LFP amplitude despite being of the same magnitude (at 30% of peak firing rate) as shown by event-triggered averages in LFP amplitude (Fig. 4e). FS cessation was associated with a larger LFP amplitude than that associated with pausing even amongst cells exhibiting both events (right-sided Wilcoxon signed rank test; n=12, p<10^−4^). Similar results were obtained indicative of significant and widespread overexcitation underlying the cessation of activity in RS units, with 225 out of 331 (68%) RS units with sufficient rate at the time of cessation exhibiting a positive value (Extended Data Fig. 7e; n=331; R_mean,cess_=0.30±0.03 [sem]; greater than 0 with p<10^−7^ by bootstrap mean resampling test, n_sample_=20, N_bootstrap_=50,000). However, amongst RS cells exhibiting a pause in activity, membrane potential appeared much more heterogenous despite controlling for firing rate at this time, appearing in a bimodal distribution amongst both inhibition and depolarization block that was not significantly different from 0 (Extended Data Fig. 7e&f; n=130; R_mean,cess_=-0.08±0.06 [sem]; greater than 0 with p=0.3782, n_sample_=10, N_bootstrap_=50,000). This may be because RS cells receive a greater diversity of input magnitudes during seizure progression than do FS cells but succumb to the same, more uniform mechanism underlying depolarization block at firing rate cessation and seizure transition, which we hypothesize is due to increasing potassium concentrations in the extracellular environment shared by all of these cells.

This sequence of significant hyperpolarization in FS cells followed by large and widespread overexcitation in all cells suggests a consistent membrane potential-based mechanism by which local cortical circuits fight but ultimately succumb to seizure progression. The dynamics in firing rate and action potential amplitude that characterize this mechanism were accurately reproduced in a computational model of the cell membrane incorporating voltage-dependent sodium and potassium conductances (Hodgkin-Huxley formalism) with stochastic background synaptic input. The model reproduced all of the observed mutual dynamics between action potential rate and amplitude during inhibitory (hyperpolarizing) pauses versus depolarization-induced cessation (Fig. 5), confirming the plausibility of this mechanistic sequence driving unit activity patterns during seizure progression.

**Figure 5.**
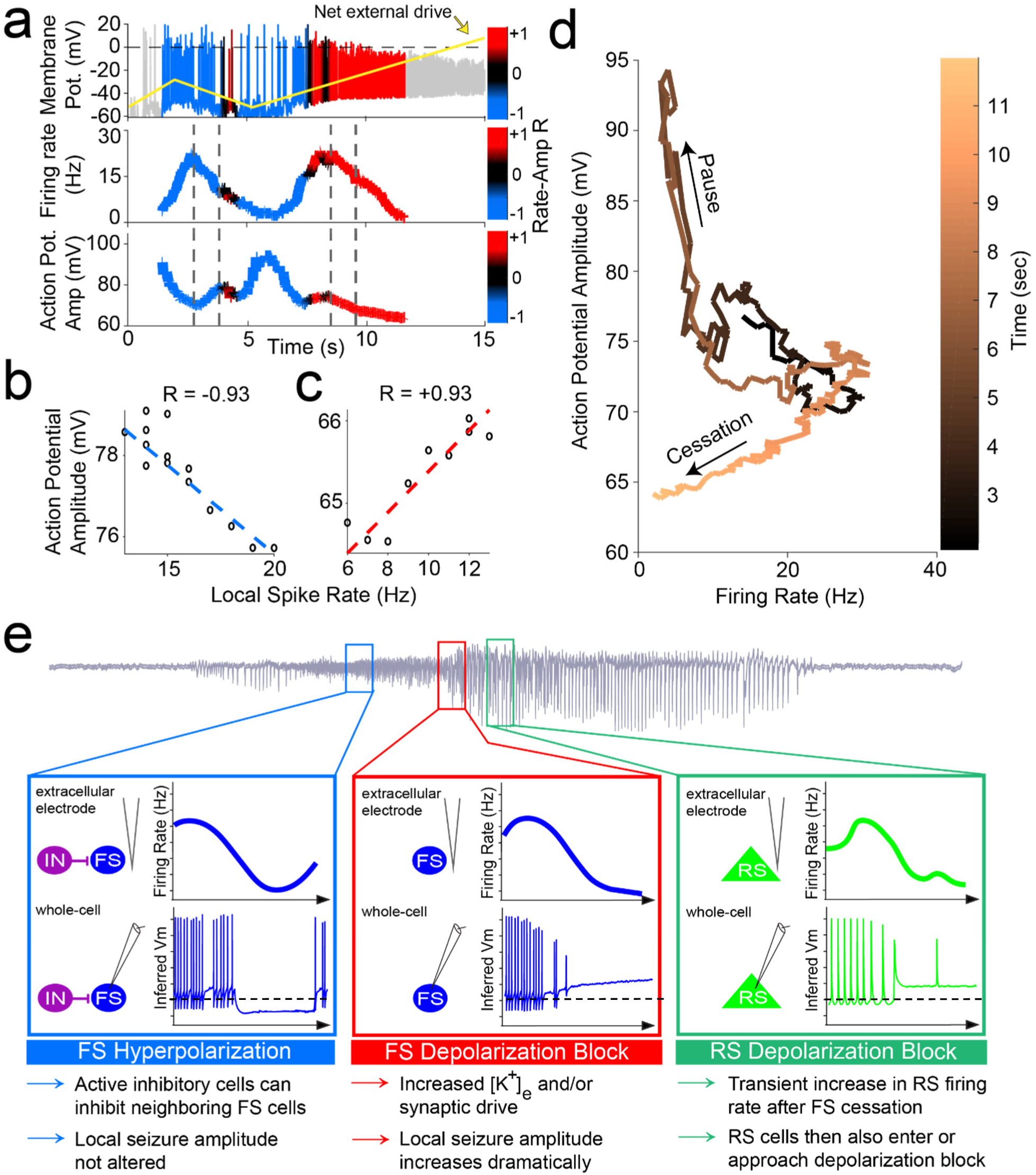
Mechanistic model of unit dynamics across seizure progression: inhibition followed by over-excitation in conductance-based neuron model reproduces key features of extracellular unit behavior during seizures. **a**. Simulated membrane potential in conductance-based spiking model color-coded by the local correlation between spike rate and amplitude and with net excitatory input indicated by gold line *(upper panel)*. Shows what is hypothesized to be happening within cortical cells during seizure progression as inferred from extracellular spike shape and rate dynamics. Firing rate *(middle panel)* and trough-to-peak spike amplitude (*lower panel*) of the same model, color-coded by the local correlation between spike rate and amplitude as an extracellular proxy for membrane potential and subthreshold input history. Dotted lines indicate starts and ends of two time periods of firing rate suppression characterized by different membrane potential signatures further characterized in (b) and (c). **b**. Example of negative correlation between local spike rate and amplitude during firing pause in (a), corresponding to activity shutdown via inhibition. Least squares linear fit indicated in dotted lines following color scheme in (a) with Pearson’s correlation coefficient indicated above plot. **c**. Example of positive correlation between local spike rate and amplitude during firing cessation, corresponding to activity shutdown via over-excitation. Least squares linear fit indicated in dotted lines following color scheme in (a) with Pearson’s correlation coefficient indicated above plot. **d**. Trajectory of unit activity over time during seizure in local spike rate vs spike amplitude space, with increasing time indicated by increasingly lighter copper color. The first time period of firing rate reduction in (a) is indicated with an arrow as “Pause” and the second time period of firing rate reduction is indicated with an arrow as “Cessation.” **e. Summary of observed dynamics in cell-type specific firing rate and inferred membrane potential trajectories**. **Blue box:** Following seizure onset, some FS cells can stop firing for brief periods (as observed on extracellular electrodes, upper panel). This pause in firing is coupled with signs of hyperpolarization in the inferred membrane potential (lower panel). **Red box:** As the seizure starts to transition to a dramatically higher amplitude FS cells stop firing (as observed on extracellular electrodes, upper panel). This cessation is coupled with signs of excessive depolarization in the inferred membrane potential (lower panel; resting membrane potential indicated as dashed line). This is likely due to elevated extracellular potassium ion concentrations by this point of the seizure or increased synaptic drive. **Green box:** The subsequent stage of high amplitude spike-and-wave rhythms corresponds to a transient increase and then decrease of firing rate amongst RS cells (as observed on extracellular electrodes, upper panel). This decrease in firing rate is coupled with signs of excessive depolarization in the inferred membrane potential (lower panel).

## DISCUSSION

We have shown that both FS and RS cells are strongly activated during the initial spread of secondarily generalized seizures. The immediate increase in RS rate at seizure onset, in particular, clearly indicates spread of the seizure, and not a failed seizure in what has been called an ictal penumbra, where no initial increases in RS rate appear^32^. Importantly, we have shown that both FS and RS cells can sometimes pause their activity due to hyperpolarization (likely due to inhibition-mediated hyperpolarization from other nearby inhibitory neurons) but stop firing midway through secondarily generalized seizures due to depolarization block. This result is independent of seizure etiology (cortical dysplasia vs. mesial temporal sclerosis) or site of origin (hippocampus vs. neocortex). This suggests that there is an almost complete lack of FS-mediated inhibition towards the end of a human seizure, in contrast to some animal models without loss of inhibition^18^. FS cells are active near the beginning and middle of the seizure, in contrast to computational models placing the loss of inhibitory restraint at seizure onset^37^. Strikingly, this absence of FS inhibitory activity is accompanied by large increases in the amplitude of the seizure’s LFP and unimpeded fast traveling waves, as well as a short-lasting increase in RS firing. Thus, these results suggest a more complicated set of dynamics than a monotonic increase or decrease of inhibition and provide further insight into the associated biophysical mechanisms.

The main difference between RS and FS cell firing during the seizures observed here was the significantly earlier cessation of spiking by FS cells, which is consistent with previous results from *in vitro* models of seizure-like events showing increased propensity of FS cells to enter depolarization block compared to pyramidal cells^12,14,25^. The reason for this difference in propensity to enter depolarization block has not been established, but these results suggest this difference could represent a primary source of imbalance between excitatory and inhibitory cell activities that allows seizures to propagate and transition to a large amplitude phase in human neocortex. One hypothesis accounting for this difference is that of the differential expression of voltage-gated potassium channel units in FS as opposed to RS cells^38–40^ in combination with the elevated extracellular potassium ion levels known to occur in the context of seizures^41,42^. However, there are several intrinsic and network-based properties that are known to differ between FS cells and pyramidal cells and could contribute to this difference, such as intrinsic excitability and the relative strength of feedforward drive^43–46^. The results here suggest that properties responsible for increasing the propensity of FS cells to enter depolarization block relative to RS cells are also those that allow the propagation and exacerbation of seizures in human neocortex. Beyond seizure progression, it is also possible that the lack of FS activity sets in motion a critical network-level transition that leads to seizure termination^47–49^. This transition may include the activation of inhibitory cell types other than FS cells^22,59^.

In summary, FS cells are the largest source of inhibition in the neocortex, and human FS cells approach cessation less than halfway through a seizure, most likely due to depolarization block. This is associated with a dramatic increase of the seizure’s local field potential amplitude and a transition to clear spike-and-wave events at a frequency of ∼3 Hz. Rhythmic spike-and-wave discharges, when they occur in the motor cortex, are responsible for the rhythmic ∼3 Hz movements seen during the clonic phase of tonic-clonic seizures^50^. Importantly, this suggests a novel, FS-dependent, mechanistic explanation for the two behaviorally defined phases of secondarily generalized tonic-clonic seizures: the high-FS-rate, low-LFP-amplitude phase of the seizure corresponds to the tonic phase, whereas the post-FS-cessation phase with periodic spike-wave bursts corresponds to a longer lasting clonic phase. Thus, in addition to existing approaches^51–55^, novel therapies that prevent FS cells from entering depolarization block may prevent - or at least limit the severity of - seizures, representing a novel and potentially powerful avenue for treating seizures with many different etiologies.

## METHODS

### Patients & Clinical/Research Electrode Placement

Approval for all experiments was granted by the Institutional Review Boards of Massachusetts General Hospital / Brigham & Women’s Hospital and Rhode Island Hospital. The decision to implant intracranial electrodes in an epilepsy patient as well the positioning of those electrodes in a patient was made solely on clinical factors by clinical staff. The explicit goal of this study was to examine the single neuron correlates of tonic-clonic seizures^42^ with focal onsets (also referred to as focal seizures with secondary generalization), one of the most common kind of epilepsies^56^. These focal seizures have a localized onset zone, either in the mesial temporal lobe or neocortex, but secondarily generalize, spreading to both hemispheres and almost always leading to impaired consciousness. Here, we studied data from 4 patients (Patients A,B,C,D) with clear focal seizures with secondary generalization, with each of these clinical seizures showing clear spike-and-wave patterns. Two additional patients with temporal-lobe epilepsy were implanted with NeuroPort Arrays but not studied here because one of them did not have any seizures while implanted (Patient E), and the NeuroPort Array in the other patient did not sample any typical spike-and-wave seizures (Patient F). A total of 10 secondarily generalized seizures with spike-and-wave discharges from the 4 patients were analyzed (3 each from Patients A and D; 2 each from Patients B and C). A detailed clinical description of each of these 4 patients follows:

#### Patient A

Patient A was a left-handed man in his 30s at the time of his surgery. He had suffered from pharmacologically intractable partial complex seizures for almost two decades. His seizures lasted 1-2 minutes and were characterized by a sudden onset of slurred and nonsensical speech. This was followed by a staring spell, lack of responsiveness, and head turning to the right. He also displayed automatisms and posturing that involved his right arm and hand more than his left. MRI suggested left (dominant) temporal polymicrogyria. He underwent placement of grids and strips for ∼2 weeks to delineate the seizure focus with respect to this area of abnormal sulcation. A 4×4 mm NeuroPort array with 1.5 mm long contacts was placed in the left superior temporal gyrus. Seizures were found to emanate from the mesial temporal structures, but during secondary generalization, the seizures spread to the location of the array in the superior temporal gyrus and beyond. The patient underwent a left temporal lobectomy, and histology confirmed that the microarray targeted layer 3. Pathology was consistent with mesial temporal sclerosis.

#### Patient B

Patient B was a right-handed man in his 40s at the time of his surgery, with a history of medically refractory epilepsy. His seizures lasted 1-2 minutes. Clinically, the seizures started with arousal and bilateral arm and leg extension. This was followed by leftward head deviation, left arm flexion, and generalized tonic-clonic activity. He underwent placement of grids, strips, and depths in his right hemisphere. A 4×4 mm NeuroPort array (with 1.5 mm deep contacts) was placed in the right middle temporal gyrus. During secondary generalization, the seizures spread to the location of the NeuroPort array in the middle temporal gyrus and beyond. The patient underwent a right temporal lobectomy. Histology on the resected tissue confirmed that the microarray targeted layer 3.

#### Patient C

Patient C was a left-handed woman in her 20s at the time of her surgery. She started to have complex partial seizures at least 10 years prior to surgery. These seizures included an aura of nausea and a ‘tunneling’ sensation, then a flattening of affect, slowed responsiveness, automatisms, and associated amnesia. Such seizures occurred 3-4 times per month and were persistent despite being on a three-drug anticonvulsant regimen. MRI revealed an extensive nodular gray matter heterotopia in the right hemisphere. Video-EEG monitoring had found right hemispheric onset seizures, and fMRI had shown normal left sided language activation patterns and normal motor activation patterns. Wada testing confirmed left hemispheric language dominance and suggested her left hemisphere could adequately support memory function subsequent to a right temporal lobectomy. Based on these data, she was implanted with a combination of subdural grid and strip electrodes over the right hemisphere and 3 depth electrodes into the right temporal lobe. The NeuroPort array (with 1.5 mm deep contacts) was placed in the right middle temporal gyrus. Her seizures lasted between 1-2.5 minutes. These showed very consistent patterns of seizure onset and propagation on ECoG; all began in the right middle and upper gyri of the posterior temporal cortex. Low amplitude and fast activity were recorded from these grid sites at the onset, followed by a buildup of 11-13 Hz activities from these leads which then spread anteriorly and inferiorly along the grid. Rhythmic spike-wave discharges were also detected soon after seizure onset spreading to several locations, including the location of the NeuroPort array in the right middle temporal gyrus. The patient underwent an extensive right temporal resection with extension posteriorly toward the right temporo-occipital junction but sparing of much of the mesial temporal structures (including the hippocampus). Histology confirmed that the array targeted layers 2/3. Pathology revealed subtle neuronal dysgenesis and focal superficial gliosis and encephalomalacia in the posterior temporal neocortex, including the recorded seizure-onset zone. Anterior temporal cortex showed mild gliosis.

#### Patient D

Patient D was a right-handed man in his 20s whose seizures began when he was a teenager. His seizures were characterized by a blank stare and oral automatisms accompanied by stiffening and posturing of the right hand. His MRI was unremarkable, but his semiology and phase I EEG recordings suggested a left temporal seizure onset. Consequently, he was implanted with several strip electrodes covering the left frontal and temporal regions. The NeuroPort array (1.0 mm deep contacts) was placed in the middle temporal gyrus about 1–2 cm posterior to the temporal tip. All of the seizures had similar clinical and electrographic signatures with a left gaze preference at onset followed by tonic and then clonic movements of the right arm. Electrographically, the seizures began with a generalized burst of sharp waves followed by sharp wave complexes that were maximal in mesial temporal leads. The participant underwent a left temporal lobectomy. Histological examination of the tissue revealed mild dysplastic changes in the lateral temporal neocortex and gliosis and moderate neuronal loss in regions CA4 and CA3 of the hippocampus.

### NeuroPort Recordings, Data Collection & Analysis

A Neuroport array (manufactured by Blackrock Microsystems) is a 10×10 grid of electrodes with an inter-electrode spacing of 400 um, giving a total size of 4×4 mm. The depth of the electrodes used in this study was either 1.5 mm (Patients A,B,C) or 1.0 mm (Patient D). 96 of the 100 electrodes were used to record the activity of individual neurons as well as the local field potential (LFP). The 0.3-7500 Hz wideband analog signal from each electrode was referenced to a distant intracranial reference wire and sampled at 30 kHz. The LFP shown in Figs. 1, 2, and Extended Data Fig. 1 were z-scored, but not filtered any further. Traveling wave analysis & images (Fig. 3) made use of these z-scored, unfiltered LFP signals. For single unit analysis, the broadband signal was high-pass filtered above 250 Hz using a 6-pole Bessel filter and then manually spike-sorted using Plexon Offline Sorter. We then classified the neurons as either fast-spiking (FS) inhibitory interneurons or regular-spiking (RS) excitatory cells using well-established criteria^27,28^, including action potential shape. Cessation of firing during a seizure was defined as the bin after which the firing rate never exceeded 30% of the unit’s peak rate. For action-potential (AP) amplitude analysis (Fig. 4a), the AP amplitude was defined as the trough-to-peak amplitude of each individual extracellular action potential.

### Cluster isolation quality assessment: Dynamic L_Ratio_

To assess the isolation quality of feature space clusters over the course of seizures, we employed a modified version of the L_ratio_ metric of cluster separation introduced and validated by Schmitzer-Torbert et al. (2005)^58^. This involved first calculating the Mahalanobis distance of every spike to a given cluster’s center in a 4D feature space consisting of trough-to-peak amplitude, trough-to-peak width, principal component 1, and principal component 2. The Mahalanobis distance normalizes the Euclidean distance by the variance of a given cluster along its major dimensions to correct for correlations amongst features. Schmitzer-Torbert et al. (2005)^58^ showed strong error rate correlation with ground truth extracellular spikes by assuming a multi-dimensional Gaussian distribution for a given cluster and taking the amount of contamination by false negative assignments to be the sum of probabilities of each un-clustered spike of a given Mahalanobis distance to belong to that Gaussian, which was robust to the particular feature space used. These probabilities are given by the inverse of the cumulative distribution function of a chi-squared distribution with degrees of freedom equal to the number of features in the features space, the sum of which is called L. Because clusters in this space moved over the course of the seizure in a non-monotonic pattern here, we calculated Mahalanobis distances to a surrogate cluster for multiple subsets of the whole cluster at 8 equal divisions over the course of the seizure and 3 divisions of the 10 minute period that occurred two minutes after the end of the seizure. A composite cluster across initial divisions was used to provide a liberal estimate of the space that a given cluster might occupy despite early waveform shape changes. This composite reference cluster included all available spikes before the seizure along with those in the first two divisions of the seizure, which were generally well-sortable. We calculated the sum of false negative assignment probabilities under a Gaussian model for this reference cluster, the value L, for each seizure and post-seizure division. We then dynamically normalized L by the number of spikes assigned to the cluster in each time division to estimate contamination rates relative to the number of spikes assigned during that division, which we refer to as the dynamic L_ratio_.

### Membrane potential trajectory analysis

We exploited the biophysical relationship between depolarization, voltage-gated Na+ channel inactivation, and spike waveform shape^60^ to infer the direction of membrane potential changes underlying the changes in firing rate exhibited by single units recorded extracellularly in patients across time. This was done for a given unit by first computing the local spike rate and average trough-to-peak waveform amplitude in moving time windows of width 1 second unless otherwise specified, moving with a step size of 0.1 seconds. Then a linear regression was performed at each point in this series across a moving time window of 3 seconds, producing a time series of correlation coefficients for bins with an average spike rate of at least 5 Hz. Strong negative correlations were taken as evidence of firing rate changes associated with membrane potential changes near resting membrane potential, while strong positive correlations were taken as evidence of membrane potential changes closer to firing threshold, near the regime of depolarization block^56^.

### Computational model

Computer simulations of transmembrane voltage dynamics consistent with the extracellular observations here were made in MATLAB R2017b to explore plausible underlying biophysical mechanisms. A 4-dimensional ordinary differential equation model of a patch of neural membrane following the Hodgkin-Huxley formalism of passive (g_L_=0.033 mS/cm^2^, E_L_=-60 mV, I_L_=g_L_(*V*-E_L_)) and voltage-dependent spiking Na+ (g_Na_=60.0 mS/cm^2^, E_Na_=+58 mV, I_Na_=g_Na_m^3^h(*V*-E_Na_)) and K+ (g_K_=5 mS/cm^2^, E_K_=-85 mV, I_K_=g_K_n^4^(*V*-E_K_)) conductances was solved using Euler integration with a timestep of 0.005 msec. First order kinetics of gating variables *m, h*, and *n* were modeled as in previously established models of this form^57^. Slowly changing net synaptic input and extracellular ion gradient changes were considered to constitute a net external drive modeled as an injected inward current. This current follows a temporal form hypothesized to drive human neocortical neurons during secondarily generalized seizures based on the dynamic membrane potential decoding analysis performed here. Fast synaptic inputs were modeled as conductances with a maximum conductance g_syn_=0.15 mS/cm^2^ and an exponential decay time course of 5 milliseconds gated by input spike times generated by a Poisson process with a linearly increasing rate parameter from 0.001 to 0.2 Hz representing increasing input firing rates across seizure. Analysis of the resulting spike trains was completed in the same manner as dynamic membrane potential trajectory decoding was performed for the observed extracellular data.

### Statistical analyses

In performing hypothesis testing on various metrics describing activity amongst FS and RS units including peak firing rate difference, mean cessation time difference, and mean correlation between spike firing rate and action potential amplitude at time of cessation, we employed bootstrap-based methods to quantify the uncertainty in these metrics. For peak firing rate and cessation time differences this involved combining all relevant unit measurements into a single population, randomly reshuffling RS and FS labels amongst this population, and recomputing the relevant difference metrics N_bootstrap_ times as indicated to generate a distribution of the metric for random unit classifications. The probability of a Type-I (false positive) error, the p-value, was then reported as the area under the distribution corresponding to values more extreme than the observed value for the original data. For testing the positivity of the mean correlation between spike firing rate and action potential amplitude at time of cessation, this involved first removing correlation coefficients in the time series corresponding to a linear regressions involving spike rates of less than 5 Hz. Then the coefficients at each unit’s detected cessation time were resampled with replacement in 20 unit subpopulations 50,000 times (unless otherwise specified) and averaged to generate a bootstrapped sample mean distribution characterizing the variability in this mean relative to zero. The probability of a Type-I (false positive) error that the observed mean coefficient was greater than zero by chance, the p-value, was then reported as the area under the distribution corresponding to values equal to or less than zero. To nonparametrically test the significance of differences in paired measures at cessation and pause times within single units, we employed the right-sided Wilcoxon signed rank test (RMatlab2017b).

## Data Availability

The seizure data that support the findings of this study are available on request from the authors. The data are not publicly available due to them containing information that could compromise research participant privacy/consent.

## Extended Data / Supplementary Information

9 Extended Data Figures (+ legends) are included.

## ACKNOWLEDGEMENTS

We would like to thank the patient volunteers. This work was supported by the American Epilepsy Society Junior Investigator Research Award, CURE Epilepsy Taking Flight Award, NINDS F32-NS083208 and University of Michigan Startup Funds (OJA), NIH R01-NS062092 (SSC) and NSF graduate student fellowships (TTJ & EKWB).

## SUPPLEMENTARY INFORMATION

**Extended Data Figure 1:**
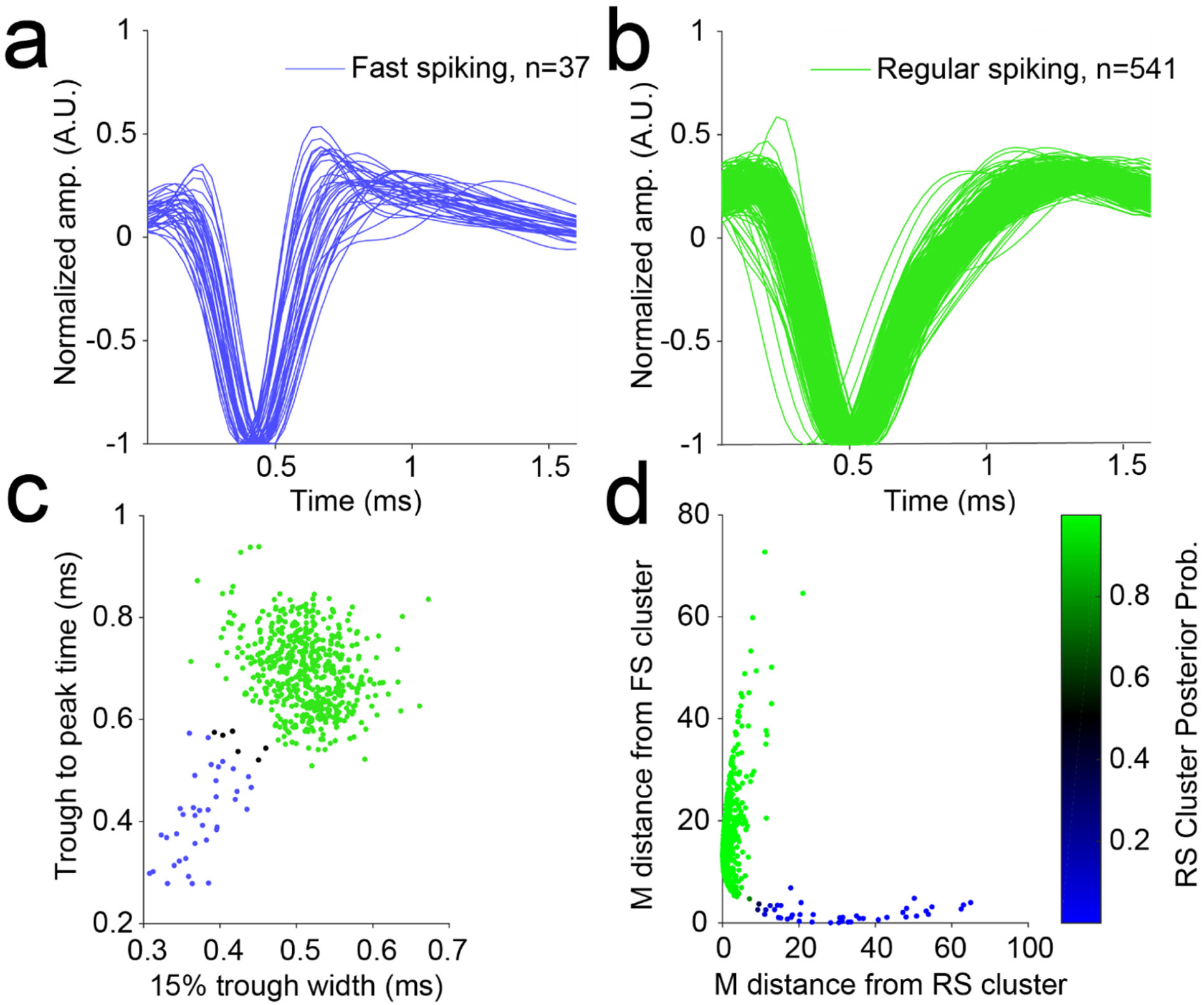
Human neural activity classification. **a**. Average waveform of each FS-classified unit, normalized by the value the waveform reaches at its trough. The overlaid average waveforms show visual consistency amongst all units classified as FS and as having spike widths narrower than those of RS-classified units. **b**. Average waveform of each RS-classified unit, normalized by the value the waveform reaches at its trough. The overlaid average waveforms show visual consistency amongst all units classified as RS and as having spike widths wider than those of FS-classified units. **c**. All units indicated as points in the average waveform feature space that produced optimal cluster separation, namely the width of the waveform at the potential corresponding to 15% of the potential at its trough and the time between the trough and peak of the waveform. Clustering was automated using a Gaussian Mixture Model where points with posterior probability exceeding 0.95 were assigned to the closest centroid. Black dots indicate units that did not exceed this threshold for either Gaussian cloud and were thus unclassified and not used in the remainder of the study. **d**. Mahalanobis distance of each unit from both the RS and FS clusters with color indicating its posterior probability of belonging to the RS cluster. Closeness of points to both axes shows that the clusters are well-separated when accounting for their different variances along different directions in feature space.

**Extended Data Figure 2:**
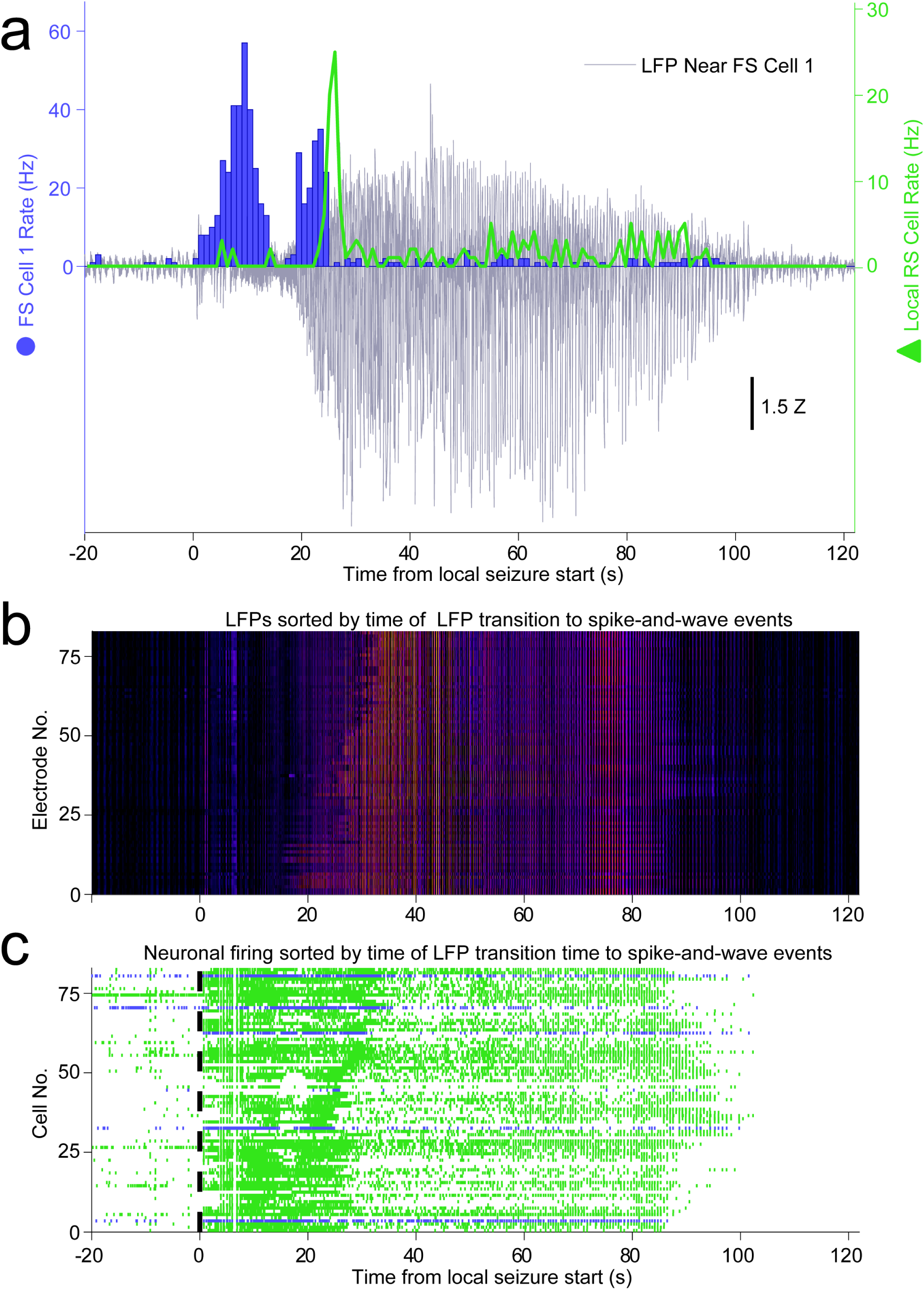
Human neocortical inhibitory and excitatory neuron activities have distinct temporal profiles relative to secondarily generalized focal seizure progression on local electrodes, Patient A, Seizure 2. **a**. The LFP (gray) recorded at the same location as FS Cell 1 (blue) dramatically increases at the same time as FS Cell 1 decreases firing. FS Cell 1 activity cessation again precedes a sharp increase in local RS cell activity (green), further suggesting an important role of FS cells in controlling local activity during seizure progression. **b**. Heatmap shows local LFP amplitude (absolute value) over time on each electrode in NeuroPort array exhibiting classifiable units as each row, sorted by time of start of spike-and-wave event and with brighter colors indicating larger amplitudes. **c**. Raster plot showing spike times of all cells on NeuroPort array in Patient B that could be classified into FS (blue) or RS (green) categories with rows sorted by the same order as in (B). Note the increasing delay to reduction in spike density corresponding to LFP transition to spike-and-wave events suggesting control of local seizure progression by local cellular spiking activity.

**Extended Data Figure 3:**
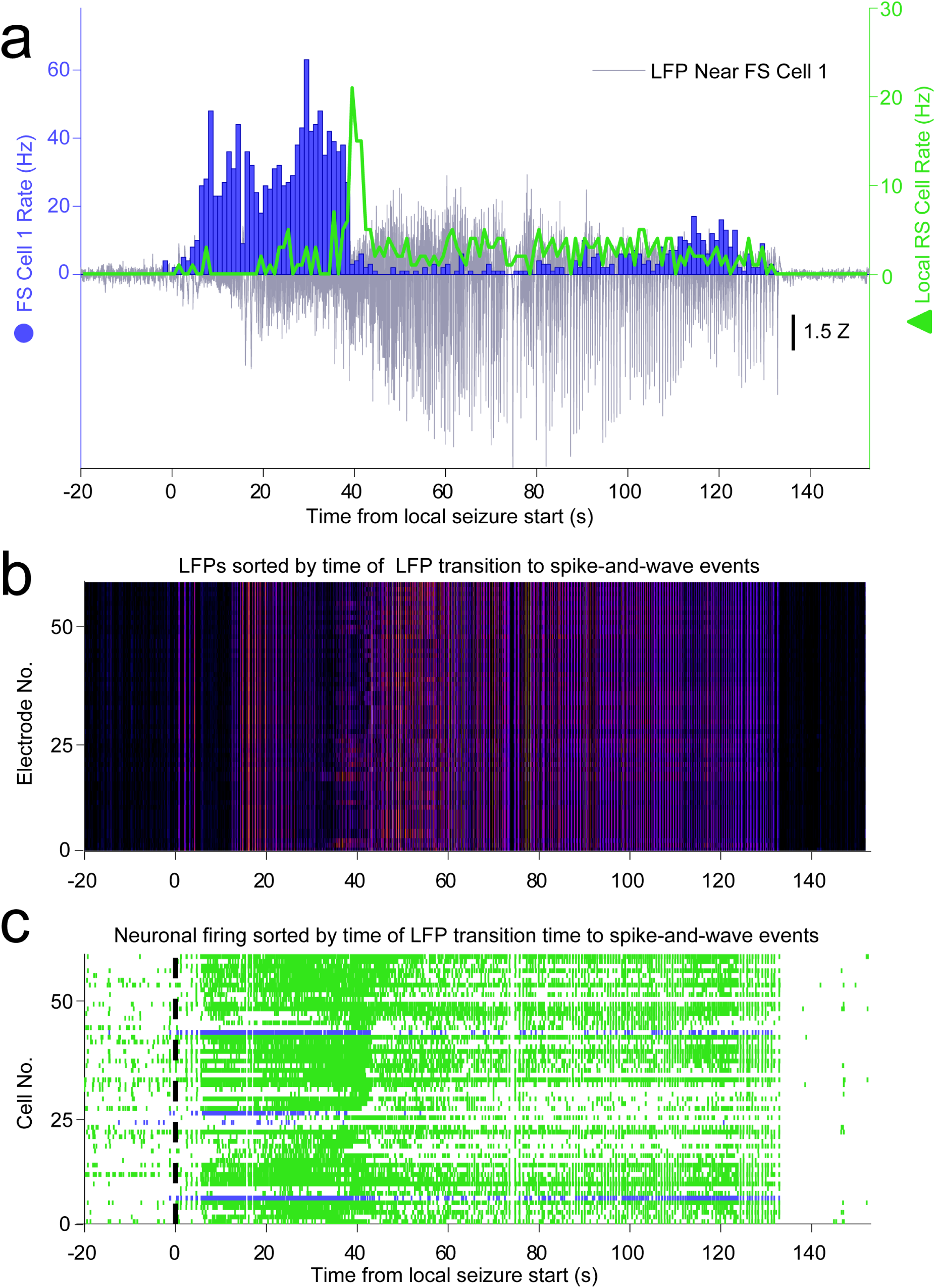
Human neocortical inhibitory and excitatory neuron activities have distinct temporal profiles relative to secondarily generalized focal seizure progression on local electrodes, Patient C, Seizure 2. **a**. The LFP (gray) recorded at the same location as FS Cell 1 (blue) dramatically increases at the same time as FS Cell 1 decreases firing. FS Cell 1 activity cessation again precedes a sharp increase in local RS cell activity (green), further suggesting an important role of FS cells in controlling local activity during seizure progression. **b**. Heatmap shows local LFP amplitude (absolute value) over time on each electrode in NeuroPort array exhibiting classifiable units as each row, sorted by time of start of spike-and-wave event and with brighter colors indicating larger amplitudes. **c**. Raster plot showing spike times of all cells on NeuroPort array in Patient C that could be classified into FS (blue) or RS (green) categories with rows sorted by the same order as in (B). Note the increasing delay to reduction in spike density corresponding to LFP transition to spike-and-wave events suggesting control of local seizure progression by local cellular spiking activity.

**Extended Data Figure 4:**
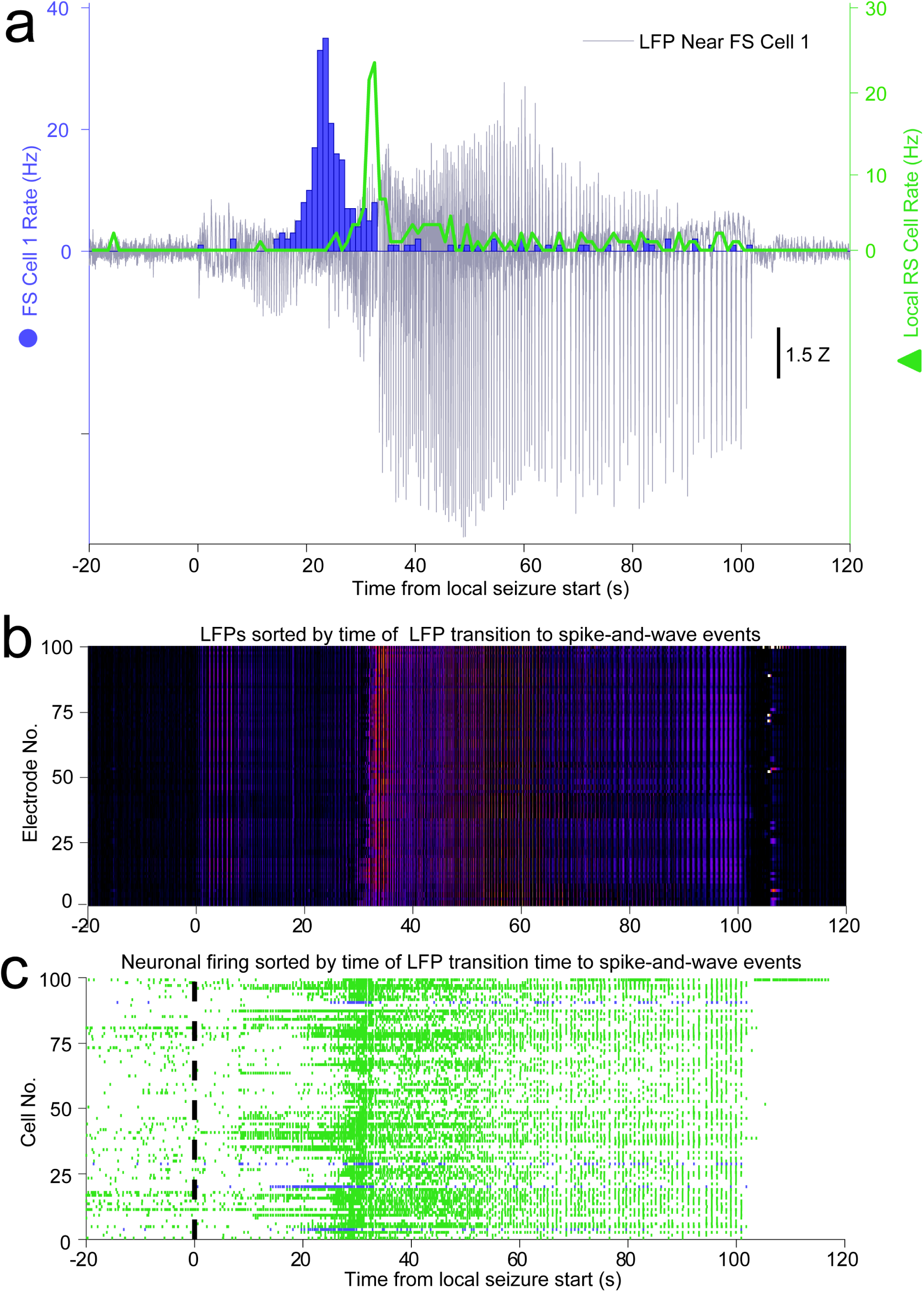
Human neocortical inhibitory and excitatory neuron activities have distinct temporal profiles relative to secondarily generalized focal seizure progression on local electrodes, Patient B, Seizure 1. **a**. The LFP (gray) recorded at the same location as FS Cell 1 (blue) dramatically increases at the same time as FS Cell 1 decreases firing. FS Cell 1 activity cessation again precedes a sharp increase in local RS cell activity (green), further suggesting an important role of FS cells in controlling local activity during seizure progression. **b**. Heatmap shows local LFP amplitude (absolute value) over time on each electrode in NeuroPort array exhibiting classifiable units as each row, sorted by time of start of spike-and-wave event and with brighter colors indicating larger amplitudes. **c**. Raster plot showing spike times of all cells on NeuroPort array in Patient C that could be classified into FS (blue) or RS (green) categories with rows sorted by the same order as in (B).

**Extended Data Figure 5:**
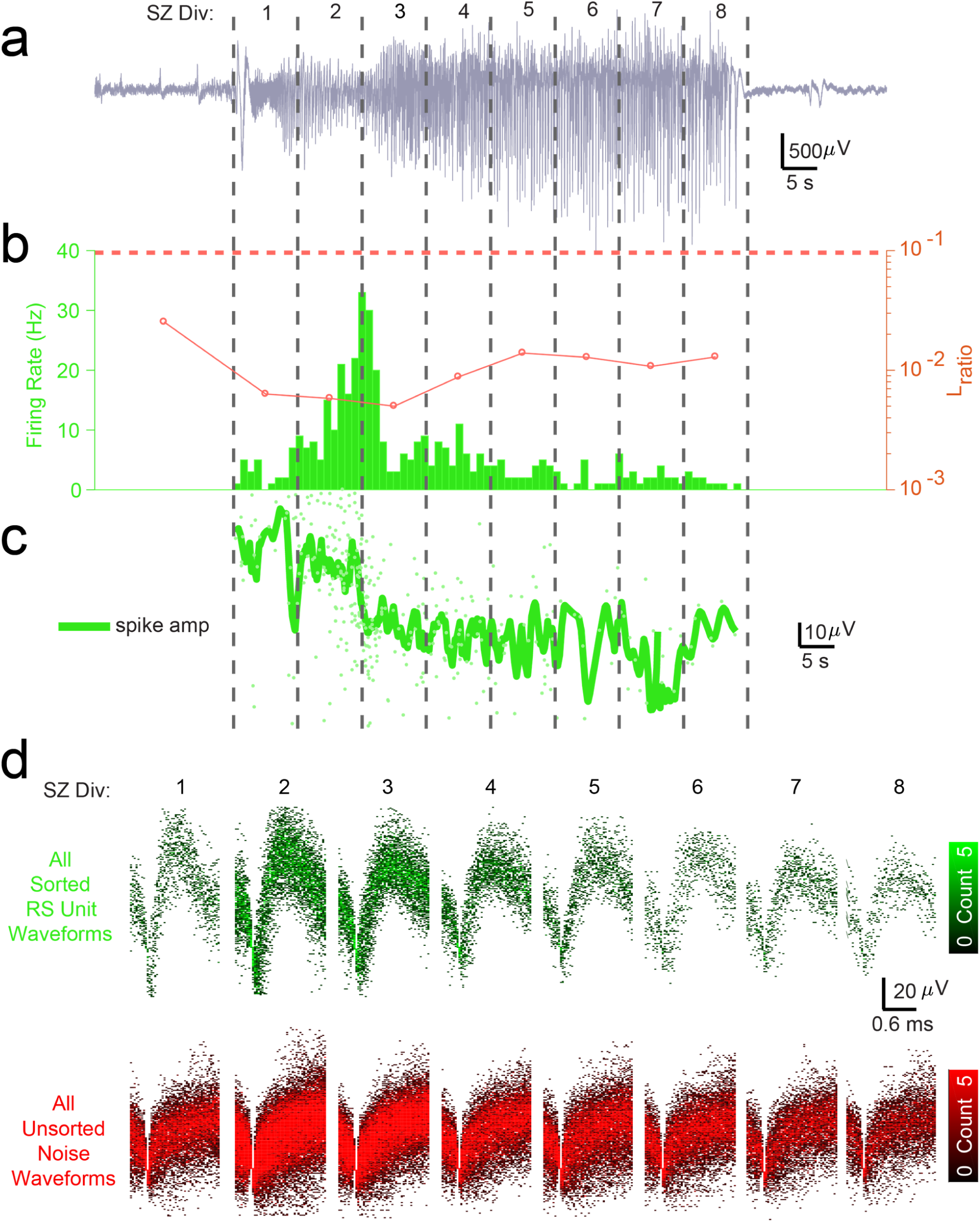
Cluster isolation quality assessment suggests RS units are well-isolatable across seizure despite changing unit amplitude and noise structure, allowing analysis of firing rate changes across seizure. **a**. LFP in Patient C indicating seizure as split into 8 equal time divisions for analysis of unit isolation quality across duration of seizure. **b**. Bar graph shows firing rate in 1 second bins of best-isolated example RS unit (green) in Patient C. Dotted red line indicates threshold used to determine best-isolated units using the dynamic L_ratio_ measure (see Methods) in each time division of seizure. Line plot indicates dynamic L_ratio_ in each division and shows large separation of example FS unit from noise in feature space used for clustering throughout the seizure (note log scale). **c**. Line plot showing average spike amplitude (dark green) and individual spike amplitudes (light green) of example RS unit over course of seizure in Patient C. Note that even as amplitude decreases the unit remains well-isolated from noise as quantified by dynamic L_ratio_ across seizure. **d**. Time-voltage histogram of all threshold crossings assigned to this example RS unit (green, *Upper)* and to noise (red, *Lower*) in eight divisions of seizure in Patient C. Shows unit waveforms are visually distinguishable from threshold crossings assigned as noise across seizure.

**Extended Data Figure 6:**
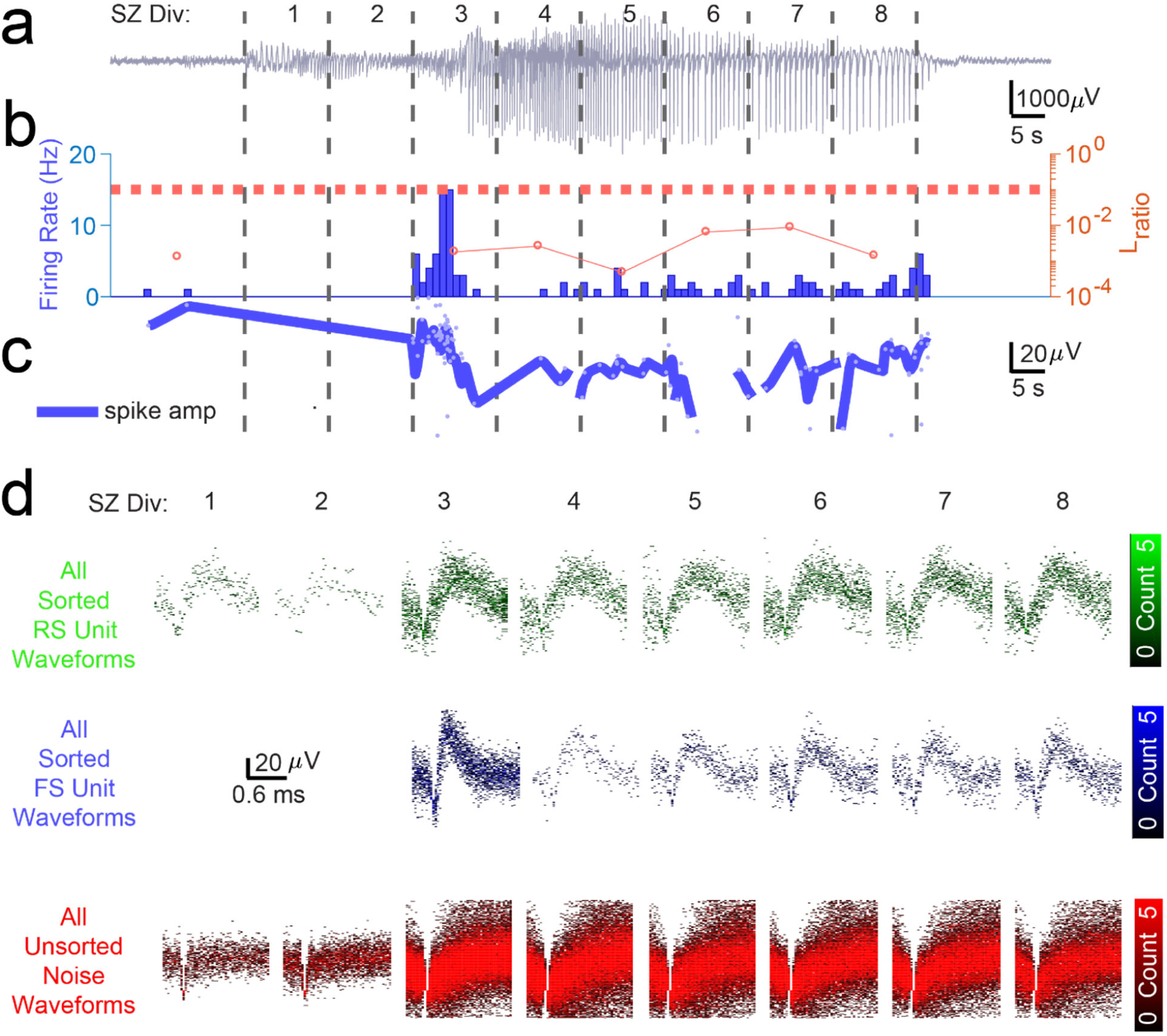
Cluster isolation quality assessment suggests RS and FS units on same channel are well-isolatable across seizure despite changing unit amplitude and noise structure, allowing analysis of firing rate changes across seizure for multiple patients. **a**. LFP in Patient B indicating seizure as split into 8 equal time divisions for analysis of unit isolation quality across duration of seizure. **b**. Bar graph shows firing rate in 1 second bins of best-isolated example FS unit (blue) in Patient B. Dotted red line indicates threshold used to determine best-isolated units using the dynamic L_ratio_ measure (see Methods) in each time division of seizure. Line plot indicates dynamic L_ratio_ in each division and shows large separation of example FS unit from noise in feature space used for clustering throughout the seizure (note log scale). **c**. Line plot showing average spike amplitude (dark blue) and individual spike amplitudes (light blue) of example FS unit over course of seizure in Patient B. Note that even as amplitude decreases the unit remains well-isolated from noise as quantified by dynamic L_ratio_ across seizure. **d**. Time-voltage histogram of all threshold crossings assigned to example FS unit (blue, *Middle)*, RS unit on same channel (green, *Upper*), and to noise (red, *Lower*) in eight divisions of seizure in Patient B. Shows unit waveforms are visually distinguishable from threshold crossings assigned as noise and as other units across seizure.

**Extended Data Figure 7:**
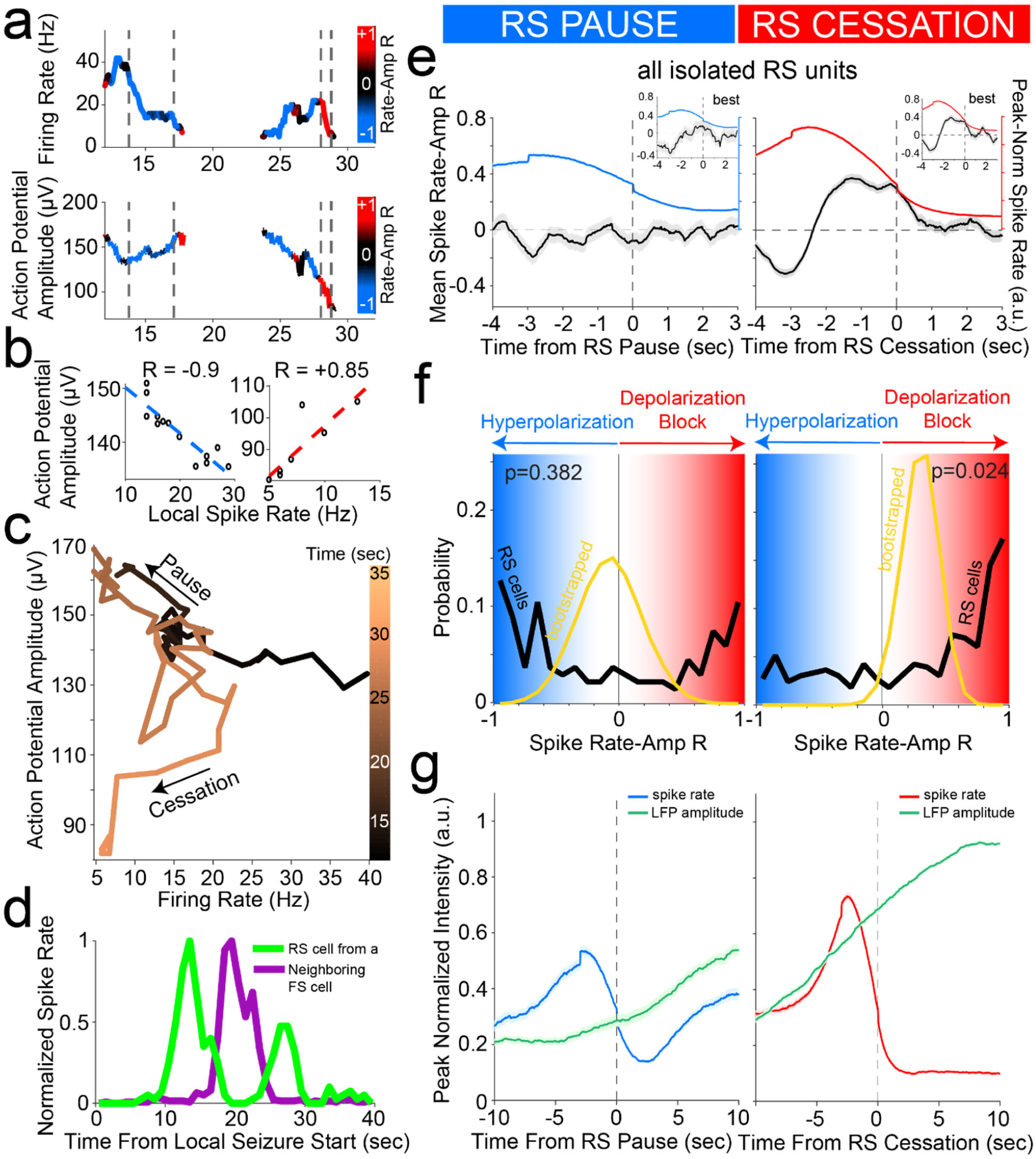
Cessation of individual RS unit activity is consistently associated with inferred membrane potential signatures of depolarization block, despite prior pauses. **a**. Firing rate *(upper)* and trough-to-peak spike amplitude (*lower*) of example RS unit in Patient A, color-coded by the local correlation between spike rate and amplitude (in 1 second time bins) as an extracellular proxy for membrane potential trajectory and subthreshold input history. Dotted lines indicate starts and ends of two time periods of firing rate suppression characterized by different membrane potential signatures further characterized in (B), namely a negative correlation regime corresponding to inhibition followed by a positive correlation regime corresponding to over-excitation ending in firing rate cessation putatively though depolarization block. **b**. Example of negative correlation (*left*) between local spike rate and amplitude in first time period indicated by dotted lines in (A) and example of positive correlation (*right*) between local spike rate and amplitude in second time period indicated by dotted lines in (A). Least squares linear fit indicated in dotted lines following color scheme in (A) with Pearson’s correlation coefficient indicated above each plot. **c**. Trajectory of unit activity over time during seizure in local spike rate vs spike amplitude space, with increasing time indicated by increasingly lighter copper color. The first time period of firing rate reduction in dotted lines in (A) is indicated with an arrow as “Pause” and the second time period of firing rate reduction in dotted lines in (A) is indicated with an arrow as “Cessation.” **d**. Firing rate of unit from (A) with neighboring FS unit firing rate overlaid, giving further evidence that first period of firing rate suppression corresponds to inhibition from local FS units while second period of firing rate suppression does not correspond to inhibition from local FS units. **e**. Unit cessation-triggered population average of the time course of novel membrane potential regime measure, i.e. the correlation coefficient relating spiking amplitude and rate in a local time window, around the two significant descents in firing rate (below 30% of peak rate) that occur in sequence during seizure. These are designated as pause (*left panel*) and cessation (*right panel*). Left panel shows population average firing rate (blue) and inferred membrane potential regime (black) aligned according to the time of pause in each unit, for all RS units displaying a pause (n=176), with inset showing average for best-isolated RS units displaying a pause (n=41). Right panel shows population average firing rate (red) and inferred membrane potential regime (black) aligned according to the cessation time of each unit, for all RS units meeting cessation criteria (n=379), with inset showing average for best-isolated RS units (n=111). **f**. Probability distribution of membrane potential regime measure (spike rate-amplitude correlation; black line) at the time of pause (*left panel*) for RS unit pausing subpopulation (n=176) with sample mean distribution (gold line; resampling size *n*=10, *N*_*bootstrap*_=50,000) showing a bimodal distribution of correlation coefficients near +1 and -1, i.e. in the hyperpolarized membrane potential regime (blue) or in the highly depolarized regime (red). The population distribution of inferred membrane potential regime is also shown at the time of cessation (*right panel*) for the RS unit population meeting cessation criteria (n=379) with sample mean distribution (gold line; resampling size *n*=10, *N*_*bootstrap*_=50,000) showing the mean correlation to be significantly above zero, i.e. in the highly depolarized membrane potential regime (red). This is indicative of widespread depolarization block occurring across RS population at the time of cessation. **g**. Unit cessation-triggered population average of the time course of same-electrode LFP amplitude around unit activity pause (*left panel*) and cessation (*right panel*). In particular, left panel shows population average firing rate (blue) and LFP amplitude (black) aligned according to the time of pause in each unit, for all RS units displaying a pause (n=176). Right panel shows population average firing rate (red) and LFP amplitude (black) aligned according to the time of cessation in each unit, for all RS units (n=379).

**Extended Data Figure 8:**
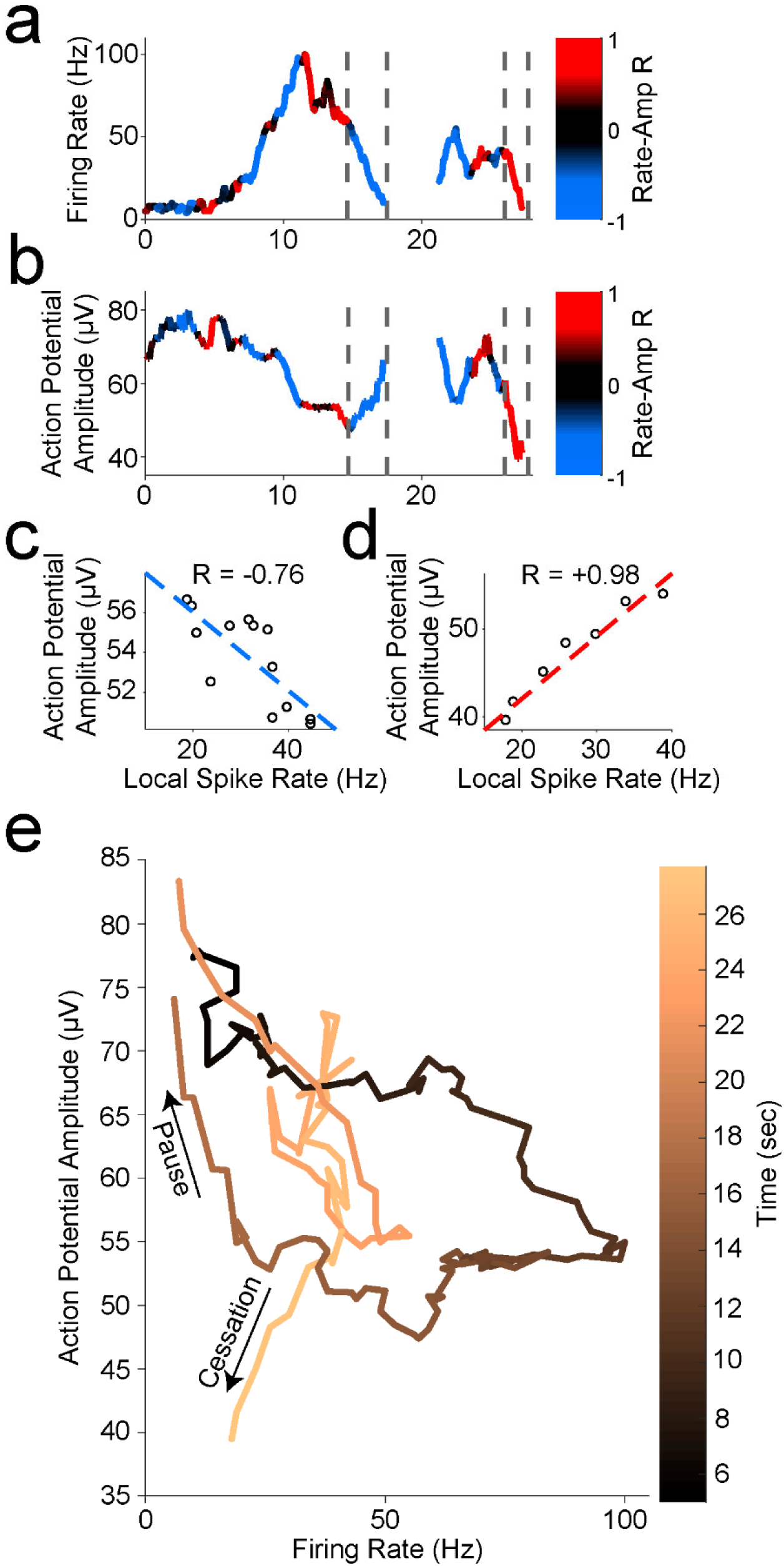
Seizure progression is consistently associated with extracellular signs of depolarization block preceded by inhibition in single units. **a**. Firing rate *(upper)* and trough-to-peak spike amplitude (*lower*) of another example FS unit in Patient A, color-coded by the local correlation between spike rate and amplitude (in 1 second time bins) as an extracellular proxy for membrane potential and subthreshold input history. Dotted lines indicate starts and ends of two time periods of firing rate suppression characterized by different membrane potential signatures further characterized in (B), namely a negative correlation regime corresponding to inhibition followed by a positive correlation regime corresponding to over-excitation ending in firing rate cessation putatively though depolarization block. **b**. Example of negative correlation (*left*) between local spike rate and amplitude in first time period indicated by dotted lines in (A) and example of positive correlation (*right*) between local spike rate and amplitude in second time period indicated by dotted lines in (A). Least squares linear fit indicated in dotted lines following color scheme in (A) with Pearson’s correlation coefficient indicated above each plot. **c**. Trajectory of unit activity over time during seizure in local spike rate vs spike amplitude space, with increasing time indicated by increasingly lighter copper color. The first time period of firing rate reduction in dotted lines in (A) is indicated with an arrow as “Pause” and the second time period of firing rate reduction in dotted lines in (A) is indicated with an arrow as “Cessation.”

**Extended Data Figure 9:**
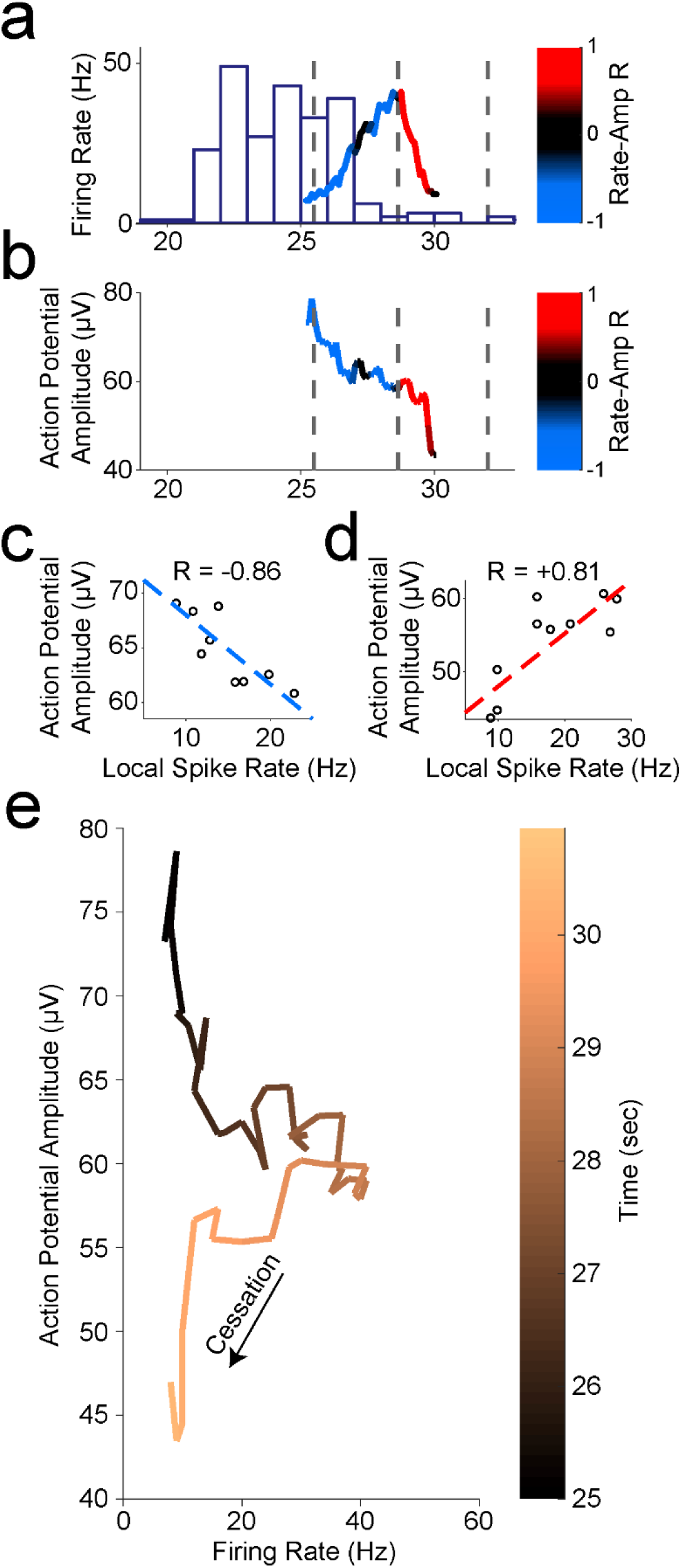
Seizure progression is consistently associated with extracellular signs of depolarization block preceded by inhibition in single units. **a**. Firing rate *(upper)* and trough-to-peak spike amplitude (*lower*) of another example RS unit in Patient A, color-coded by the local correlation between spike rate and amplitude (in 1 second time bins) as an extracellular proxy for membrane potential and subthreshold input history. Firing rate of local FS unit is overlaid as purple histogram. Dotted lines indicate start and end times of two time periods of firing rate suppression characterized by different membrane potential signatures further characterized in (B), namely a negative correlation regime corresponding to inhibition followed by a positive correlation regime corresponding to over-excitation ending in firing rate cessation putatively though depolarization block. **b**. Example of negative correlation (*left*) between local spike rate and amplitude in first time period indicated by dotted lines in (A) and example of positive correlation (*right*) between local spike rate and amplitude in second time period indicated by dotted lines in (A). Least squares linear fit indicated in dotted lines following color scheme in (A) with Pearson’s correlation coefficient indicated above each plot. **c**. Trajectory of unit activity over time during seizure in local spike rate vs spike amplitude space, with increasing time indicated by increasingly lighter copper color. RS unit is released from inhibition shortly following the cessation of activity in local FS unit as indicated by increasing firing rate with decreasing amplitude but then enters regime of over-excitation indicated by decreasing firing rate with decreasing amplitude at trajectory “corner” corresponding to a spiking rate of 40 Hz, until the unit ceases to fire likely due to depolarization block given these indicators of membrane potential history.

